# Strong anti-viral responses in pediatric COVID-19 patients in South Brazil

**DOI:** 10.1101/2021.04.13.21255139

**Authors:** Tiago Fazolo, Karina Lima, Julia C. Fontoura, Priscila Oliveira de Souza, Gabriel Hilario, Renata Zorzetto, Luiz Rodrigues Júnior, Veridiane Maria Pscheidt, Jayme de Castilhos Ferreira Neto, Alisson F. Haubert, Izza Gambin, Aline C. Oliveira, Raissa S. Mello, Matheus de Bastos Balbe e Gutierres, Rodrigo Benedetti Gassen, Ivaine Tais Sauthier Sartor, Gabriela Oliveira Zavaglia, Ingrid Rodrigues Fernandes, Fernanda Hammes Varela, Márcia Polese-Bonatto, Thiago J. Borges, Sidia Maria Callegari-Jacques, Marcela Santos Correa da Costa, Jaqueline de Araujo Schwartz, Marcelo Comerlato Scotta, Renato T. Stein, Cristina Bonorino

## Abstract

Epidemiological evidence that COVID-19 manifests as a milder disease in children compared to adults has been reported by numerous studies, but the mechanisms underlying this phenomenon have not been characterized. It is still unclear how frequently children get infected, and/or generate immune responses to SARS-CoV-2. We have performed immune profiling of pediatric and adult COVID-19 patients in Brazil, producing over 38 thousand data points, asking if cellular or humoral immune responses could help explain milder disease in children. In this study, pediatric COVID-19 patients presented high viral titers. Though their non-specific immune profile was dominated by naive, non-activated lymphocytes, their dendritic cells expressed high levels of HLA-DR and were low in CX3CR1, indicating competence to generate immune responses that are not targeted to inflamed tissue. Finally, children formed strong specific antibody and T cell responses for viral structural proteins. Children’s T cell responses differed from adults in that their CD8+ TNFα+ T cell responses were low for S peptide but significantly higher against N and M peptide pools. Altogether, our data support a scenario in which SARS-CoV-2 infected children may contribute to transmission, though generating strong and differential responses to the virus that might associate with protection in pediatric COVID-19 presentation.

## Introduction

COVID-19 is a complex disease, with multisystemic involvement, and an array of clinical manifestations that can vary from asymptomatic to severe outcomes leading to death (1) which lead to an ongoing worldwide emergency (2). Epidemiological evidence of less severe forms of the disease and reduced mortality in children upon infection with SARS-CoV-2 has been consistently reported (3,4), except for an inflammatory syndrome (MISC) associated with co-morbidities in a relatively low percentage of children (5). The pediatric population (0-19 years old) represents more than 25% of the Brazilian population, however, it is observed that this group corresponds to only 1.9% (19,589 / 989,170) of all cases of SARS by COVID-19 reported in the past 12 months. Mortality among children represented 0.5% (1,564 / 321,659) of all deaths due to the disease reported in the same period. The lethality in children and adolescents hospitalized due to SARS by covid-19 was 8.0% (1,574 / 19,589), while the overall lethality in all age groups was 32.5% (321,659 / 989,170), in the observed period (data from SIVEP-Gripe, Ministry of Health). Thus, a significantly lower number of children and adolescents has severe clinical presentations with the need for hospitalization, or that will lead to death, when compared to other age groups.

Different hypotheses have been raised to explain this phenomenon (6,7). Milder disease in children could result from a reduced expression of the viral receptor ACE2, leading to lower levels of viral replication (8). Alternatively, it could be explained by a differential immune response in children, correlated to a distinct infection course from adults (9). A third popular hypothesis is that the pre-existence of neutralizing antibodies to seasonal coronaviruses could confer some cross-protection against SARS-CoV-2 induced disease, mainly because children are considered the main reservoir for these viruses (10). At present, the scarcity of data prevents a clear understanding of the striking differences between the pediatric and adult outcomes after infection by COVID-19.

Comprehensive studies have characterized immune responses in adults with mild or severe forms of COVID-19 (11–14). However, considerably fewer studies have focused on pediatric patients. This is a subject of paramount importance, not only because it is central to the design of public policies regulating school opening (and all the activities associated with it) during the pandemic, but also because understanding the milder disease presentation in children may provide important clues for the design of prevention strategies as well as novel therapeutic pathways for the management of COVID-19. In this study, we sought to characterize in detail the innate and adaptive immune responses in a cohort of patients consisting of adults with different degrees of severity and children with mild disease presenting at health care facilities with symptoms suggestive of COVID-19. We sought to identify an immune profile in children that could explain the striking differences in outcome between them and adult COVID-19 patients. We collected plasma and peripheral blood mononuclear cells (PBMCs) from adult and pediatric COVID-19 patients, and detailed characterization of their immune response was performed by multi-parameter flow cytometry, defining 78 immune cell subsets and expression of key activation markers, anti-SARS-CoV-2 IgA and IgG antibodies, and frequencies of specific effector T cells, producing 38,670 data points. Pediatric patients with mild COVID-19 had high viral load titers, high frequencies of dendritic cells, with high HLA-DR expression, but low in CX3CR1. Although their non-specific adaptive cells immune profile was dominated by antigen-inexperienced cells, SARS-CoV-2 specific antibodies and T cells were detected in levels comparable to the ones in adults with either severe or mild disease. Children showed higher CD8+ TNFα+ T cell responses to N and M peptide pools than for S peptides, while this was not observed in adults. This response did not correlate with anti-S or anti-N antibody levels. Taken together, our findings suggest that children produce a differential immune response when compared to adults, which associates with the mild manifestation in pediatric COVID-19.

## Methods

### Ethics Statement

This study was approved by the Institutional Review Board (IRB 30749720.4.1001.5330) at Hospital Moinhos de Vento and by the Ethics Committee at Universidade Federal de Ciências da Saúde (CAAE 30749720.4.3001.5345). Legal consent was obtained from all participants or their legal guardians. The study was conducted according to good laboratory practices and following the Declaration of Helsinki.

### Patients

A prospective cohort study was carried out at Hospital Moinhos de Vento and at Hospital Restinga e Extremo Sul, both in Porto Alegre, southern Brazil. A convenience sample of adults and children older than 2 months were enrolled from June to December 2020 at either the outpatient clinics (OPC), emergency rooms (ER), or hospitalized. Subjects were screened if presenting cough and/or axillary temperature ≥37.8°C and/or sore throat. Both blood samples and respiratory samples collected through nasopharyngeal swabs were obtained at enrollment. Patients with the clinical diagnosis of COVID-19 and SARS-CoV-2 infection confirmed by RT-PCR were included in the study. Clinical and demographic data were collected at inclusion, following a standardized protocol. Disease severity was classified according to the World Health Organization classification after completing the follow-up questionnaire (15).

### SARS-CoV-2 RT-q-PCR

A qualitative RT-PCR assay to SARS-CoV-2 was performed for all participants. Bilateral nasopharyngeal and oropharyngeal swabs were collected and placed in the same transport medium with saline solution and RNAlater®, RNA Stabilization Solution (Catalog number AM7021, Invitrogen™). MagMax™ Viral/Pathogenic Nucleic Acid Isolation Kit (Applied Biosystems) was used to extract viral RNA in the KingFisher Duo Prime System (ThermoFisher, USA) automated platform. The RT-PCR assay was performed in 10 µL total reaction, using Path™ 1-Step RT-qPCR Master Mix, CG (catalog number A15299, AppliedBiosystems) and TaqMan™ 2019-nCoV Assay Kit v1 (catalog number A47532, AppliedBiosystems) which comprises the SARS-CoV-2–specific targets (gene ORF1ab, gene S and gene N). As reaction control was used 5 µL (200 copies/µL) the TaqMan™ 2019-nCoV Control Kit v1 (catalog number A47533, AppliedBiosystems). QuantStudio 5 (ThermoFisher Scientific, USA) was used to perform the PCR.

### PBMC isolation and cryopreservation

Blood was collected in EDTA tubes (Firstlab, PR, Brazil) and stored at room temperature before processing for PBMC isolation and plasma collection. Plasma was separated by centrifugation and cryopreserved. PBMCs were next isolated by density-gradient centrifugation using Ficoll–Paque™ PLUS (GE Healthcare®), and either studied directly or resuspended in FBS 5% DMSO and stored in liquid nitrogen until use.

### Flow cytometry

Cells were thawed by diluting them in 5mL pre-warmed complete RPMI1640 medium (Sigma-Aldrich - R8758) containing 5% FBS and spun at 1500 rpm for 5 minutes. Supernatants were carefully removed, and cells were resuspended in PBS. After, were stained with the BD Horizon™ Fixable Viability Stain 510 together with antibodies for surface markers, as follows: anti-CD3-APC-H7 (clone SK7), anti-CD24-APC-H7 (clone ML5), anti-HLA-DR-APC-H7 (clone G46-6), anti-CD4-PerCP-Cy5.5 (clone RPA-T4), anti-CD27-PerCP-Cy5.5 (clone M-T271), anti-CD11c-PerCP-Cy5.5 (clone B-ly6), anti-CD14-PerCP-Cy5.5 (clone M5E2), anti-CD8-FITC (clone HIT8a), anti-IgG-FITC (clone G18-145), Lineage 2-FITC (cat. 643397), anti-CD16-FITC (clone 3G8), anti-CXCR5 (CD185)-BB515 (clone RF8B2), anti-CD19-APC (clone HIB19), anti-CD127-Alexa 647 (clone HIL-7R-M21), anti-CX3CR1-Alexa647 (clone 2A9-1), anti-CD69-APC (clone FN50), anti-CD38-PE (clone HIT2), anti-ICOS (CD278)-PE (clone DX29), anti-CD141-PE (clone 1A4), anti-CD66b-PE (clone G10F5), anti-CD137 (4-1BB)-PE (clone 4B4-1), anti-HLA-DR-PE-Cy7 (clone G46-6), anti-CD19-PE-Cy7 (clone SJ25C1), anti-CD25-PE-Cy7 (clone 2A3), anti-CD45RA-PE-Cy7 (clone L48), anti-IgM-BV421 (clone G20-127), anti-PD-1 (CD279)-BV421 (clone MIH4), anti-CD303-BV421 (clone V24-785), anti-CD56-BV421 (clone NCAM 16), anti-CCR7-BV421 (clone 2-L1-A) antibodies. For intracellular staining, cells were first stained for surface markers and subsequently fixed and permeabilized using the Transcription Factor Buffer Set (BD Biosciences-Pharmingen, USA), then stained with anti-Ki-67-BV421 (clone B56), anti-Perforin-Alexa 647 (clone δG9), and anti-Granzyme B-BV421 (clone GB11) antibodies. Following *in vitro* stimulation assays with specific peptides, cells were stained with the BD Horizon™ Fixable Viability Stain 510 and anti-CD3-PE-Cy7 (clone SK7), anti-CD4-PerCP-Cy5.5 (clone RPA-T4), anti-CD8-APC-H7 (clone SK1), anti-CCR7-BV421 (clone 2-L1-A), and subsequently fixed and permeabilized using the Cytofix/Cytoperm kit (BD Biosciences-Pharmingen, USA), then stained with anti-IFNγ-FITC (clone 4S.B3), anti-TNF (clone MAb11) and anti-IL-17-PE (clone SCPL1362) antibodies. All samples were analyzed using a BD Biosciences - FACSCanto II and FlowJo 10.7.1 software.

### *In vitro* T cells stimulation assays

PBMC were thawed, assayed for viability, counted, and plated in 96-well plates at 3×10^6^ PBMCs/mL, 100 uL/well in RPMI1640 medium (Sigma-Aldrich - R8758) supplemented with 10% fetal bovine serum (100 IU of penicillin/mL, 100 μg of streptomycin/mL (Lonza, Belgium) and 2 mM L-glutamine (Lonza, Belgium) (R10H medium), and subsequently stimulated with peptide PepTivator SARS-CoV-2 Prot S (130-126-700 - Miltenyi Biotec, Germany), PepTivator SARS-CoV-2 Prot N (130-126-698 - Miltenyi Biotec, Germany) and PepTivator SARS-CoV-2 Prot M (130-126-702 - Miltenyi Biotec, Germany) at 1 μg/mL. PMA (50 ng/mL, Sigma, USA) plus ionomycin (1 μg/mL, Cayman chemical company, USA) and DMSO were used as positive and negative controls for stimulation, respectively. Stimulation with a CMV peptide pool at 2μg/mL (Mabtech, Sweden) was also performed, as a positive control for the assay. All treatments were submitted for 18h at 37°C and 5% CO_2_. Three hours before harvesting, Golgi Plug (BD Biosciences, USA) 1μg/mL was added to each well. Cells were stained and analyzed for phenotype as described above.

### ELISA

Plasma was tested for IgG and IgA antibodies to S-RBD protein (#RP-87678 - Invitrogen, USA) and N protein (kindly provided by Dr. Ricardo Gazinelli - Fiocruz Belo Horizonte, Brazil) using a protocol described in (16). Briefly, ELISA plates (Kasvi, Brazil) were coated overnight with 1μg/mL of SARS-CoV-2 Spike Protein (S-RBD). On the following day, plates were blocked for 1 h at room temperature with blocking buffer (3% Skim Milk Powder in Phosphate Buffered Saline (PBS) containing 0.05% Tween-20). Plasma samples were heat-inactivated at 56°C for 60 minutes and then serially diluted in 1% milk in 0.05% PBS-Tween 20 starting at a 1:25. Plasma was incubated for 2 h at 37°C. Secondary antibodies were diluted in 0.05% PBS-Tween and incubated for 1 h at room temperature. For both IgG, anti-human peroxidase produced in rabbit (#IC-1H01 - Rhea Biotec, Brazil), and IgA, anti-human peroxidase produced in goat (#A18781 - Invitrogen, USA), was used at a 1:10,000 dilution. The assay was developed with TMB Elisa Substrate - High Sensitivity (Abcam, United Kingdom) for 30 minutes, and the reaction stopped with 1M chloric acid. Readings were performed in an ELISA reader (Biochrom EZ 400), and O.D. at 450 nm was used to calculate the area under the curve (AUC), using a baseline of 0.07 for peak calculation (17).

### Statistics

Percentages were used to describe categorical variables. Pearson’s Chi-square test was used to evaluate proportions among the children, severe and mild adults. Data normality assumptions were verified for continuous variables and summarized in terms of median and interquartile range (IQR). Two-tailed Kruskal-Wallis test followed by Benjamini-Hochberg correction for multiple comparisons was used to compare values among the groups. Principal Component Analysis (PCA) was employed to reduce the dimensions of 78 immunological variables generated by flow cytometry analysis, to explain the total variability with a smaller, new set of variables. Spearman correlations were performed between all variables (every set of two variables) and within sets of variables to identify clusters of correlated variables. In PCA of the variables grouped in clusters, variables containing redundant information were excluded. Comparison among groups regarding single variables or PC values was performed by non-parametric Kruskal-Wallis test, followed by 2 by 2 multiple comparisons with p-values adjusted accordingly. All analyses were performed either in GraphPad Prism v.9 or R and sometimes confirmed in Python. 3D analysis of PCA was plotted in Python. Scripts are detailed in supplemental materials.

## Results

The study design is summarized in Figure 1A. We have recruited a total of 92 patients (25 children; 34 adults with mild disease - AMD; and 33 adults with severe disease - ASD). All subjects had COVID-19 confirmed by PCR. All children had mild disease and were treated as outpatients. Their characteristics are described in Table 1. Patients’ characteristics and mean cycle threshold (Ct) values are displayed in Table 1. As expected, comorbidities were concentrated in the group with severe disease, which was also the group with a higher mean age. Some symptoms are probably not accurately assessed in some children, such as anosmia or dysgeusia, due to the age of some individuals in this group. Median Ct levels for all three probes used in PCR were higher in AMD, and not different between ASD and children.

**Table 1.** Clinical characteristics of all patients in this study.

| Characteristics | Mild<br>(n = 34) | Severe<br>(n = 33) | Children<br>(n = 25) | P-value |
| --- | --- | --- | --- | --- |
| Age (y), median (IQR) | 37.8<br>(27.0-44.6) | 60.8<br>(38.8-75.9) | 9.0<br>(1.5-13.5) | **<0.0001\*** |
| Female sex, n (%) | 22 (64.7) | 17 (51.5) | 10 (42.0) | 0.1657† |
| Active or passive smoking, n (%) | 2 (5.9) | 6 (18.2) | 3 (12.0) | 0.0791† |
| **Racial or ethnic group** |  |  |  |  |
| Caucasian, n (%) | 28 (82.4) | 20 (60.6) | 17 (68.0) | 0.1129† |
| Non-caucasian, n (%) | 4 (11.8) | 1 (3.0) | 5 (20.0) |  |
| **Days from of symptom onset to sample collection** |  |  |  |  |
| Days, median (IQR) | 18.0<br>(16.0-20.5) | 10.0<br>(7.5-14.0) | 15.0<br>(8.5-17.5) | **<0.0001\*\*** |
| **Symptoms** |  |  |  |  |
| Headache, n (%) | 32 (94.1) | 23 (69.7) | 14 (56.0) | **0.0262†** |
| Myalgia, n (%) | 30 (88.2) | 20 (60.6) | 9 (36.0) | **0.0030†** |
| Malaise, n (%) | 28 (82.4) | 31 (93.9) | 14 (56.0) | **0.0006†** |
| Coryza, n (%) | 26 (76.5) | 18 (54.5) | 16 (64.0) | 0.2167† |
| Cough, n (%) | 25 (73.5) | 30 (90.9) | 17 (68.0) | 0.0782† |
| Fever, n (%) | 23 (67.6) | 26 (78.8) | 20 (80.0) | 0.4899† |
| Chills, n (%) | 21 (61.8) | 20 (60.6) | 9 (36.0) | 0.0823† |
| Dyspnea, n (%) | 20 (58.8) | 22 (66.7) | 4 (16.0) | **0.0003†** |
| Dysgeusia, n (%) | 20 (58.8) | 12 (36.4) | 6 (24.0) | 0.1580† |
| Sore throat, n (%) | 19 (55.9) | 12 (36.4) | 11 (44.0) | 0.2986† |
| Appetite loss, n (%) | 19 (55.9) | 21 (63.6) | 13 (52.0) | 0.6226† |
| Anosmia, n (%) | 19 (55.9) | 11 (33.3) | 6 (24.0) | 0.1841† |
| Stuffy nose, n (%) | 17 (50.0) | 11 (33.3) | 13 (52.0) | 0.3868† |
| Conjunctivitis, n (%) | 16 (47.1) | 10 (30.3) | 7 (28.0) | 0.2465† |
| Nausea, n (%) | 14 (41.2) | 12 (36.4) | 7 (28.0) | 0.7791† |
| Sputum production, n (%) | 12 (35.3) | 10 (30.3) | 6 (24.0) | 0.6420† |
| Diarrhea, n (%) | 12 (35.3) | 16 (48.5) | 7 (28.0) | 0.2122† |
| Vomiting, n (%) | 2 (5.9) | 4 (12.1) | 4 (16.0) | 0.4442† |
| Skin rash, n (%) | 1 (2.9) | 1 (3.0) | 1 (4.0) | 0.9744† |
| **Underlying medical conditions** |  |  |  |  |
| Obesity, n (%) | 10 (29.4) | 13 (39.4) | 0 (0.0) | **0.0021†** |
| Hypertension, n (%) | 6 (17.6) | 15 (45.5) | 0 (0.0) | **0.0111†** |
| Asthma, n (%) | 1 (2.9) | 2 (6.1) | 6 (24.0) | **<0.0001†** |
| Diabetes mellitus, type 1 and 2, n (%) | 1 (2.9) | 11 (33.3) | 0 (0.0) | **0.0362†** |
| Cancer, n (%) | 1 (2.9) | 2 (6.1) | 0 (0.0) | 0.7180† |
| Tuberculosis, n (%) | 0 (0.0) | 1 (3.0) | 0 (0.0) | 0.7809† |
| Stroke/CVA, n (%) | 0 (0.0) | 4 (12.1) | 0 (0.0) | 0.2168† |
| COPD, n (%) | 0 (0.0) | 4 (12.1) | 1 (4.2) | 0.4088† |
| Heart failure, n (%) | 0 (0.0) | 2 (6.1) | 0 (0.0) | 0.6926† |
| Congenital heart disease, n (%) | 0 (0.0) | 1 (3.0) | 0 (0.0) | 0.7044† |
| **Ct value (median, IQR)** |  |  |  |  |
| ORF1ab | 19.3<br>(16.7-22.6) | 24.3<br>(19.9-29.7) | 19.8<br>(15.4-27.5) | **0.0488\*\*** |
| S | 19.8<br>(16.7-23.0) | 25.3<br>(21.9-27.7) | 19.4<br>(13.2-28.3) | **0.0239\*\*** |
| N | 18.7<br>(16.1-23.5) | 24.0<br>(20.6-28.6) | 19.7<br>(14.1-29.2) | **0.0201\*\*** |
| **Oxygen use** |  |  |  |  |
| Oxygen use during hospitalization, n (%) | 0 (0.0) | 22 (66.7) | 0 (0.0) | **<0.0001†** |

IQR = interquartile range; \*\* Kruskal-Wallis test; † Pearson's Chi-squared test

**Figure 1:**
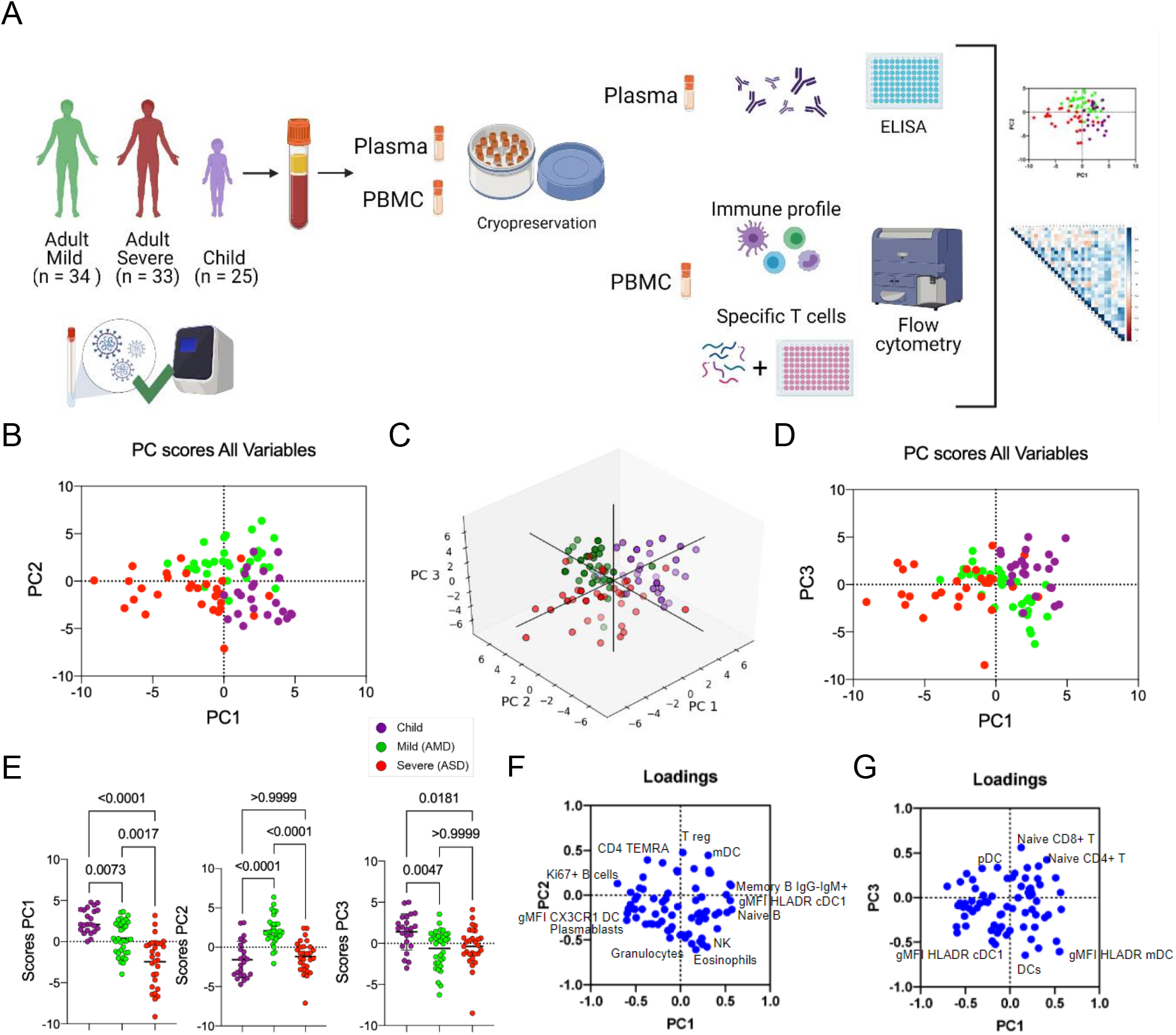
Experimental approach and differential immune profile of children, mild and adult patients by principal component analysis. **A**, Graphical representation of the study design; **B-E**, Principal component analysis of the clusters of pediatric (purple) and adult patients with mild (green) and severe (red) disease; each dot represents a patient, color coded. **B**, distribution of clusters by PC1 and PC2; **C**, 3D representation including PC1, 2 and 3; **D**, two-dimensional plot of patients according to PC3 by PC1; **E**, comparison of scores for each PC by analysis of variance (Kruskal-Wallis). **F-G**, Contribution of variables (loadings) to PC1xPC2 (**F**) and PC1xPC3 (**G**). Each blue dot is a variable. Variables with the highest contributions (negative or positive) to each PC are specified. P values are indicated over brackets.

### Immune responses separate pediatric patients, adults with mild and severe disease from each other

Comprehensive immune profiling of PBMCs from pediatric and adult patients was performed using flow cytometry, generating 78 variables (frequencies of cell subpopulations and gMFI of activation markers). Gating strategies are detailed in Supplementary Figure 1 (A-H). To reduce the dimensionality of the numerous variables, a PCA analysis was carried out. This approach indicated that the three groups (pediatric patients; mild adult patients -AMD; and severe adult patients - ASD) separated from each other based on three of the new variables created, PC1, PC2, and PC3. PC1 separated the three groups, while PC2 separated mild patients from severe ones (Figure 1B). 3D plotting performed to include PC3 showed that children again separated from mild and severe adult patients (Figure 1C) and confirmed in 2D (Figure 1D). The PC scores from all three groups were plotted and the significance of these differences was analyzed by a Kruskal-Wallis (KW) test. Children had the highest mean score value for PC1 (2.765), followed by AMD (0.142) and ASD (−2.351), and these differences were highly significant (Figure 1E). For PC2, AMD had a mean score of 2.079, higher (p<0.0001) from children and ASD (mean scores of −1.585 and −1.180, respectively, p>0.999). PC3 scores were highest in children (mean=1.384) and significantly different from adults with mild (mean=−0.629; p<0.0047) and severe (mean=−0.247; p<0.0181) disease.

Principal components are calculated based on correlations among variables, and to interpret the meaning of each PC we analyzed the positive or negative contributions (loadings) of each variable in each PC. The variables with the main positive and negative contributions (loadings) are identified in Figures 1F, for PC1 and PC2; and 1G, for PC3. Respective loadings values are listed in Supplementary Table 1. PC1 had positive inputs mainly by IgM+ memory B cells, naïve B cells, and cDC1 DR expression; and main negative contributions by proliferating B cells; plasmablasts; and CX3CR1+ expression in dendritic cells (DC) (Figure 1F). That indicated that the group with the highest mean scores for PC1 (children) would be characterized by a profile of predominantly naive or low-affinity memory B cells, not activated/differentiated; and their dendritic cells would be high in DR, but low in CX3CR1. The opposite would be true for individuals with the lowest mean scores (ASD), while AMD would be characterized by an intermediate profile for these variables. The main positive influences for PC2 were T regs, mDCs, and TEMRA cells, indicating mild adult patients present significantly higher frequencies of these cell subpopulations. The main negative influences for PC2 were eosinophils, NK cells, and granulocytes (Figure 1F), and these should be the lowest in AMD. For PC3 (Figure 1G), the main positive contributions came from naive CD8+ T cells, naive CD4+ T cells, and pDCs; and main negative contributions were DCs; and expression of HLA-DR in mDC and cDC1. Because children had the highest scores for PC3, they should have significantly higher frequencies of such cells (and higher expression of HLA-DR in DCs) compared to both AMD and ASD. To verify these PCA interpretations, we compared them to the analysis of variance (Kruskal-Wallis) results performed among the three groups of patients regarding the three most relevant, positive (Figure 2A) and negative (Figure 2B) influencers, variables for PC1, PC2, and PC3. The comparison of the three groups for their variances regarding each variable agreed with the differences among them detected by PCA. A pattern that emerged from these two combined analyses was that children, AMD and ASD, separate from each other based mostly on differences in targeting innate inflammatory responses to inflamed tissues, and the state of activation of B and T lymphocytes.

**Figure 2.**
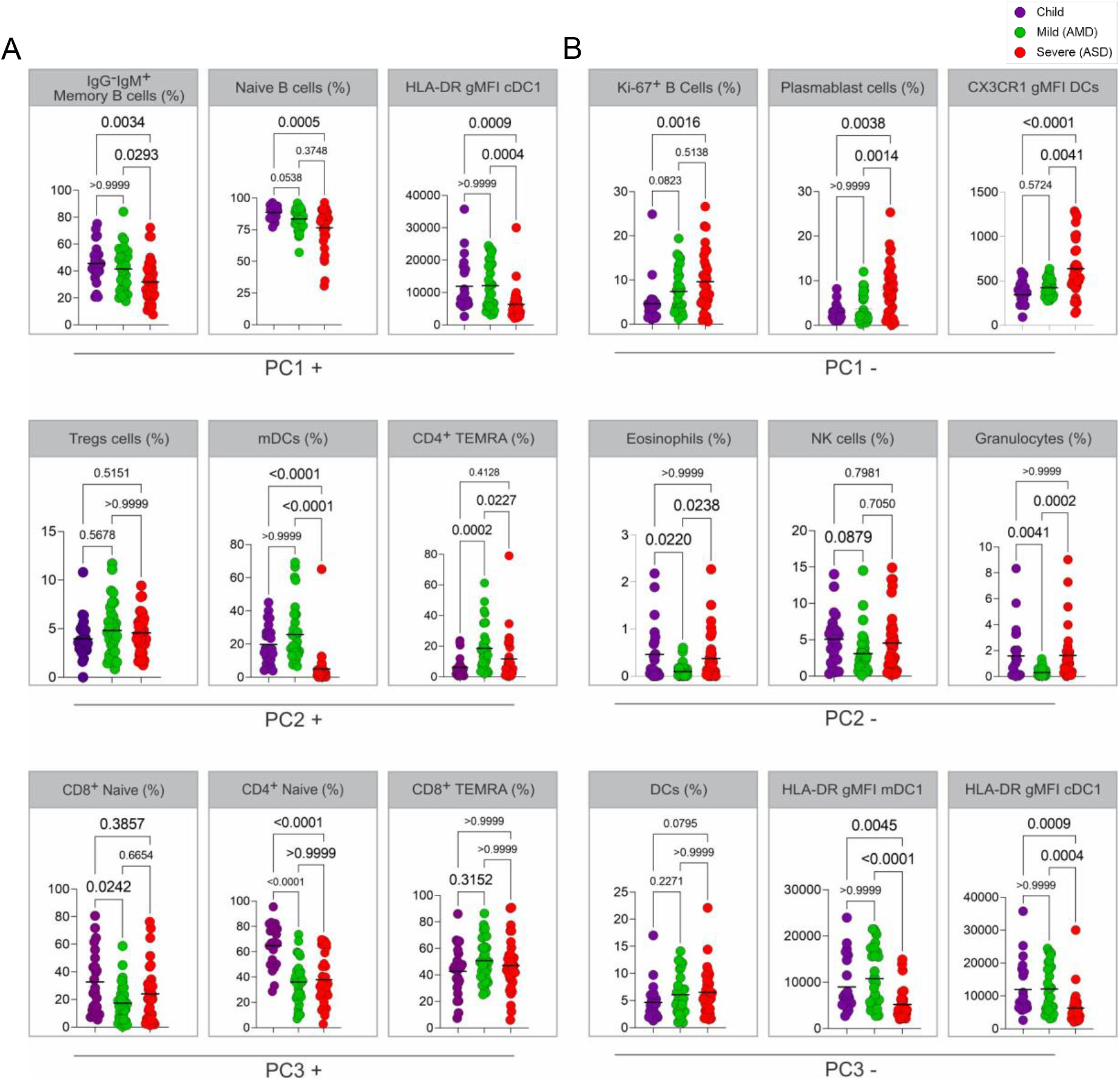
Analysis of variance of the main variables contributing to principal components. **A-B**, Kruskal-Wallis tests comparing values of each of the three immune variables that presented the highest influences – either positive (**A**) or negative (**B**) for PC1, PC2 and PC3. Each dot represents a patient, color coded: children – purple, adult with mild disease – green, and adult with severe disease – red. P values are indicated over brackets.

The percent of the total variability explained by these first principal components was low; the first three PCs together explained only 28.81% of the variance (Supplementary Table 1). This indicated that these 78 variables were not highly correlated as a whole. A correlation analysis using Spearman’ s coefficient confirmed this observation (Supplementary Figure 2), revealing a general pattern of moderate to weak correlations, but also identifying clusters of variables that were more correlated than others. These clusters represented six types of immune “signatures”: Proliferating/activated T cells; DCs; Granulocytes + Monocytes; NK cells; B cells; and memory T cells. Follicular helper T cells (Tfh) related variables were weakly correlated and were not considered as a cluster. We thus performed six separated PCAs for the identified clusters of variables, hypothesizing that they could bring more specific information to explain immunological differences among the three groups of patients. In this analysis, the first two PCs for each cluster were now explained a larger portion of the total variance (44.19, 63.17, 50.37, 67.35, 49.56, and 42.49%, respectively - Supplementary Table 1) among the three types of patients, and their distributions in children, AMD and ASD were analyzed. The results are shown in Figure 3 (A-D) and Supplementary Figure 3 (A-C), with the respective graphic representations for scores and loadings. We started by analyzing PCs formed by the innate cells’ signatures. Principal components for the clusters of Granulocytes+Monocytes (Figure 3A) and NK cells (Figure 3B), although derived from expressive correlations among their respective variables, did not separate the three groups of patients, indicating that the individuals were not significantly different for the variables that composed these PCs. That was intriguing because those variables had, as stated above, important contributions for the PC2 of all variables (Figure 1), but it also indicated that this contribution helped separate the groups mostly based on their correlations with other variables in the group of all variables, and not on the differences among groups for those variables alone. Although highly correlated among them (Supplementary Figure 2), variables composing the signatures for Granulocytes + Monocytes and NK cells did not separate the groups when used together in a PCA (not shown) and did not generate any new information.

**Figure 3.**
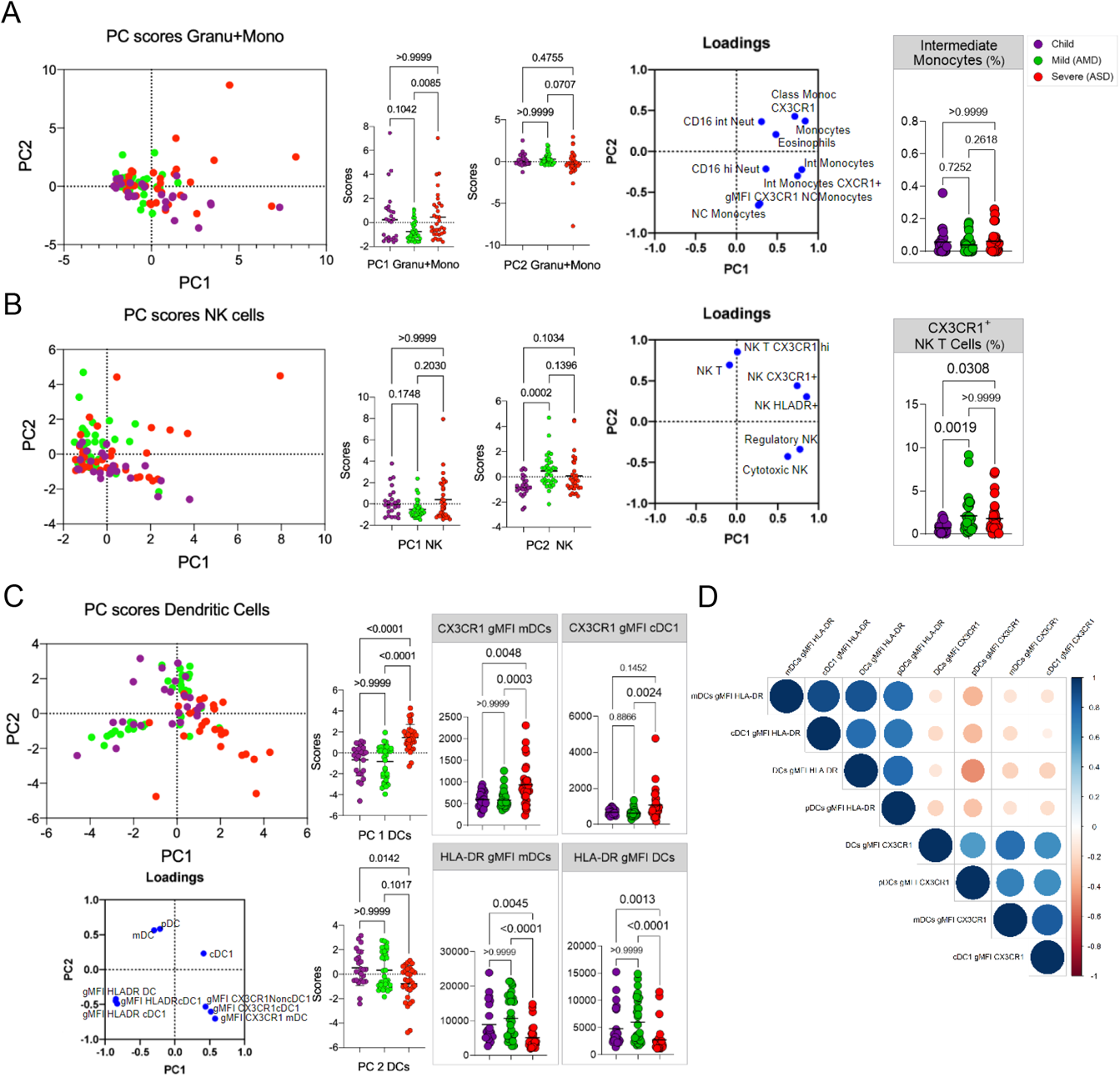
Principal Component Analysis of innate cells immune signatures. **A-G**, Principal component analysis of the clusters of patients (each dot representing a patient, color coded), according to the immune signatures (**A**, Granu^+^Mono, Granulocytes and Monocytes; **B**, NK cells; **C**, Dendritic Cells; **D**, Spearman correlation analysis of HLA-DR and CX3CR1 expression in DCS. For each signature, are displayed the PCA plot of PC1xPC2, the differences in scores of individuals for each PC; the loadings of the main variables contributing to each PC and Kruskal-Wallis tests comparisons of the major contributing variables values for each group of patients.

Findings in PCA for the DCs signature (Figure 3C) indicated that children were significantly separated from ASD, but not from AMD, with lower scores for PC1 (mostly CX3CR1 expression in DCs) and higher scores for frequencies of DCs (Figure 3C). HLA-DR expression in DCs subpopulations constituted negative contributions for PC1. This indicated that children, as well as AMD, would have more DCs than ASD, high in HLA-DR and low in CX3CR1. That was confirmed by the KW analysis of the three groups (Figure 3C). Spearman correlation analysis (Figure 3D) evidenced that CX3CR1 expression was negatively correlated with HLA-DR expression. These results suggested that high DCs frequencies in blood, with low CX3CR1, but high HLA-DR expression could be involved, or at least serve as markers, for mild disease. Inversely, low DCs frequencies, with high CX3CR1 expression, could be associated with more severe disease.

PCA for the clusters involving adaptive cells variables was performed next and are shown in Supplementary Figure 3. B cells (Supplementary Figure 3A, Supplementary Table 1) and T cell activation/proliferation (Supplementary Figure 3B, Supplementary Table 1) PCA corroborated a general pattern of response in children, either separating from the other groups (only sometimes grouping with AMD, apart from ASD). KW analysis of mean scores for each PC showed that in some cases children presented some significant differences from AMD. For example, PC1 of B cells recapitulated findings from the first analysis, mainly positively influenced for IgM+ B cells and naive cells, and showed children with significantly higher scores, compared to mild and ASD (p<0.01). Children were grouped with AMD (p=0.1599), and apart from ASD (p<0.0001), regarding PC1 of T cell activation/proliferation (Supplementary Figure 3B). The ASD had the highest scores for this PC, highly influenced by activated and proliferating CD4+ and CD8+ T cells, suggesting that adults with severe disease were characterized by higher frequencies of activated, Ki67+ T cells, and the opposite would be observed for children and AMD – though this was not always corroborated by the KW analysis. Finally, PCA for T cell memory clustered variables showed a trend to separate the three groups (Supplementary Figure 3C). PC1 scores, strongly positively influenced by naive T cells, but negatively influenced by effector memory T cells (TEM), significantly separated children from AMD (p=0.0005), and these somewhat separated from ASD (p=0.0410), indicating children and AMD would have lower frequencies of TEM and higher frequencies of naive T cells compared to ASD. Terminally differentiated memory CD4+ and CD8+ T cells (TEMRA) were strong positive influences for PC2, while expression of CD69 and CD137 in TEM cells negatively influenced PC2. Children and AMD, with high scores for PC2, did not differ from each other, suggesting they would both be characterized by higher frequencies of TEMRA cells than ASD, which in turn would have higher frequencies of activated, CD69+, CD137+, TEM cells. Confirmations of the interpretations of these PCs were again sought in the KW analysis for individual variables next to each PCA results (Supplementary Figure 3C), and also in Supplementary Figure 4 - which compiles all the remaining variables KW analyses results. This led us to note that for TEMRA, children differed from AMD and not from ASD. CD45RA, a marker upregulated both in naïve and TEMRA cells, has been shown to show different expression during the generation of memory pools of chronic infections (18) as well as in response to vaccination (19). Thus, the PCA could indicate possible differences for children in pathways for the generation of memory, although not confirmed in the KW comparisons. For TCM, there were no differences among groups (Supplementary Figure 4). Altogether, the patterns revealed by PCA indicated that children have higher frequencies of non-specific antigen inexperienced B and T cells and DCs, with high HLA-DR and low CX3CR1 expression. In some cases, children and AMD shared not only a mild presentation of the disease but also a similar immune profile. Finally, the immune profile of ASD was characterized by higher frequencies and markers of T and B cell activation and proliferation, TEM cells, and lower DCs with high expression of CX3CR1.

**Figure 4.**
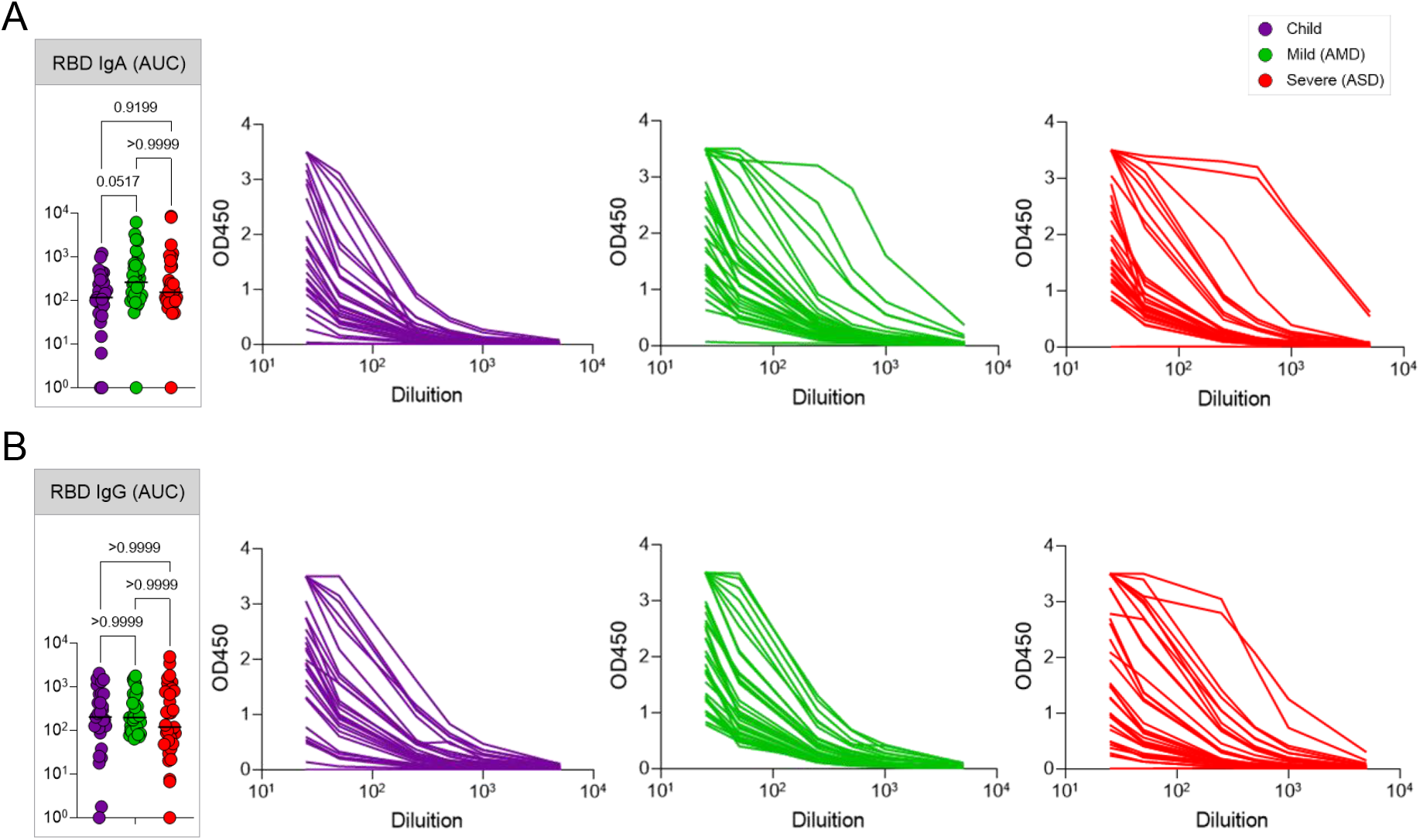
Antibody responses. SARS-CoV-2 spike RBD IgA and IgG antibody titers determined by ELISA using serial dilutions of plasma. Individual titration curves for each individual (represented by a line, color coded) and analysis of variance (Kruskal-Wallis) of the values calculated as the area under the curve (AUC) for IgA (**A**) and IgG (**B**) are displayed. P values are displayed over brackets.

### SARS-CoV-2 specific T cells and antibodies responses in children are comparable to the ones of adult patients

The characteristics of the non-specific immune profile of children led us to ask if they had effectively formed SARS-CoV-2 specific responses upon infection. Seroconversion after infection with SARS-CoV-2 patients with all forms of the disease has been described by several studies - reviewed in (20). Antibodies to the S protein, and more specifically to the RBD of this protein, are clinically considered a hallmark of infection, and frequently proposed as a correlate of protection. We thus compared children, AMD, and ASD for their RBD specific-IgA and IgG titers. On average, children presented levels of both IgG and IgA comparable to the adult patients (Figure 4). In our cohort, although some individuals from the ASD group presented higher levels of antibodies, differences among the groups were non-significant. Our results revealed that, even though children presented a generally naive, non-activated, immune profile, they had efficiently generated SARS-CoV-2 specific antibody responses, in levels that did not differ from the ones in AMD (p<0.103 for IgA; p>0.999 for IgG) or ASD (p<0.916 for IgA; p>0.999 for IgG) COVID-19 patients.

We next asked if children had generated specific effector T cell responses to SARS-CoV-2. There are four structural proteins in SARS-CoV-2: the spike glycoprotein (S), the envelope (E) protein, the membrane (M) protein, and the nucleocapsid (N) protein. Specific effector T cell responses have been described in adult COVID-19 patients, both with mild and severe disease (21,22), however fewer studies have focused on specific immune responses in pediatric patients infected with SARS-CoV-2. We measured the frequencies of CD4+T and CD8+T cells expressing TNFα, IFNγ, or IL-17 in response to stimulation by peptide pools of the S, N, and M proteins of the virus (Figure 5). Figure 5A shows representative flow cytometry plots of cytokine-producing CD4+ or CD8+ T cells upon stimulation with SARS-CoV-2 peptide pools. Negative (DMSO) and positive (PMA + ionomycin) control representative plots can be seen in Supplementary Figure 5. Children presented detectable CD4+ and CD8+ T cell responses upon stimulation with all three peptide pools (Figure 5B). When we compared types of responses in each group for the different peptide pools, children showed a significantly higher CD8+T TNFα+ response for the M (p<0.005) and for the N (p<0.0409) peptide pools than for the S pool (5B, upper right panel). This was not seen in adults, and although there was a trend for lower CD4+ TNFα+ responses in children, it was not significant. Supplementary Figure 6 shows the responses compared among the groups. About 30% of individuals – of all groups - did not show responding CD4+ T cells to the peptide pools; a higher frequency of individuals did not respond to the S pool compared to the M and N pools. In the ones that responded, CD4+ IL-17+ T cell responses were higher for all three peptide pools (about 1 log higher than INFγ and TNFα CD4+T cell responses). CD8+ T cell responses were, in general, more robust, although for S and M peptide pools there were still some individuals, though fewer, that did not respond to stimulation. The absence of response, in our sample, did not correlate with the early time of collection, as reported by (18). TNFα+ CD8+ T cell responses were about 1 log higher than what was detected for CD4+ T cells, for all peptide pools (Supplementary Figure 6). The IL-17+ CD4+T cells responses to stimulation by all three pools, were higher than the TNFα+CD4+ and INFγ+ CD4+ for all three groups. The differential TNFα+ cytotoxic response to M and N peptides seen in children led us to investigate levels of anti-N antibodies. Children made strong anti-N IgG levels, not different from AMD and ASD (Figure 6A). Anti-RBD IgA, but not IgG levels, correlated positively with CD4+ INFγ+ responses (Figure 6B). Interestingly, the TNFα+ cytotoxic responses to M and N peptide pools were inversely correlated with levels of anti-RBD and anti-N antibodies (Figure 6B). Anti-N antibody levels correlated positively with anti-N CD4+INFγ+ responses (Figure 6B). Taken together, these results indicate that children do generate specific humoral and effector cell responses upon infection with SARS-CoV-2, with a differential, higher cytotoxic response against proteins M and N, not associated with antibody responses to the spike protein.

**Figure 5.**
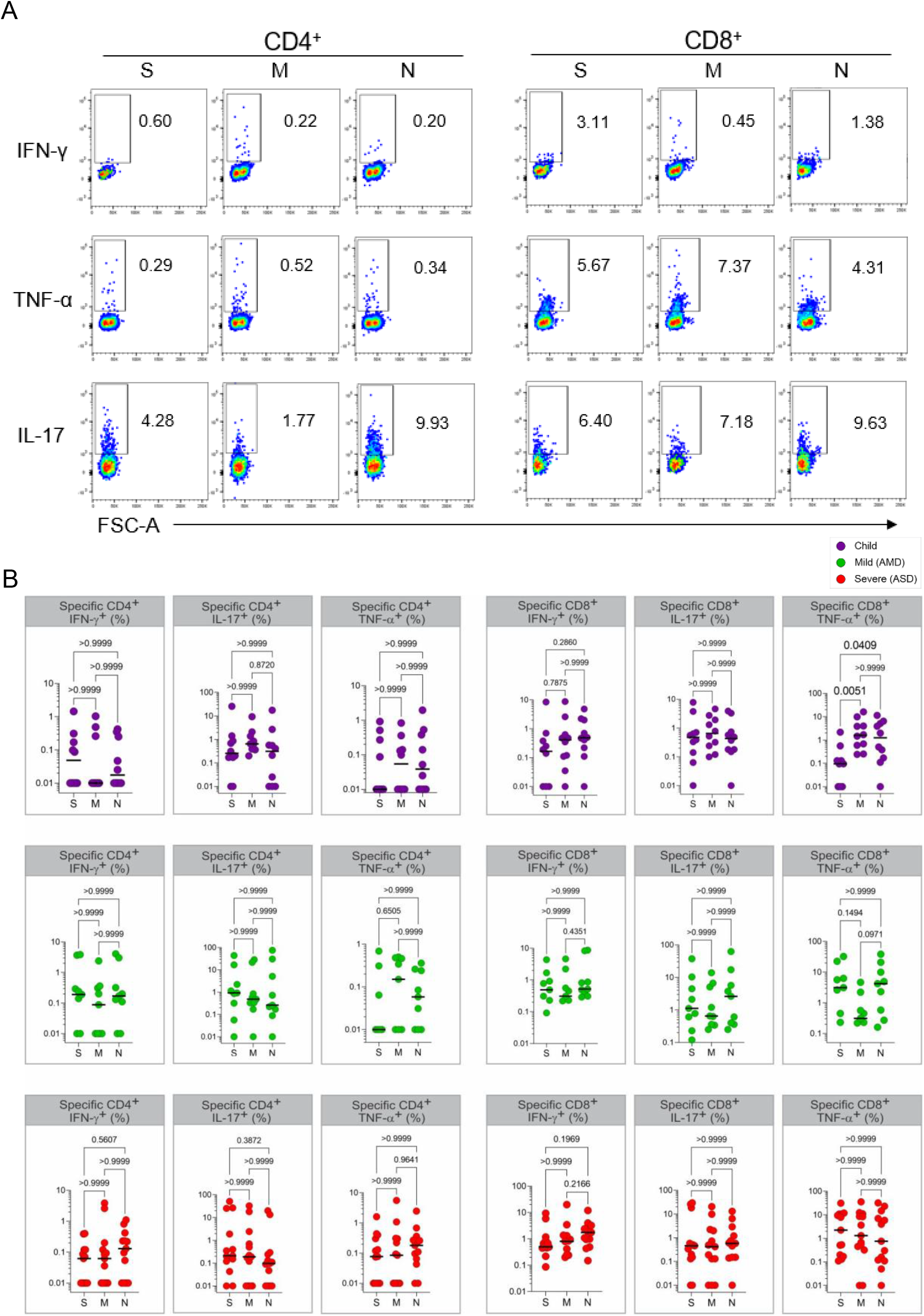
Specific T cell responses. **A**, gating strategies and typical plots of CD4+ and CD8+ T cells stimulated with peptide pools from structural proteins spike (S), membrane (M) and nucleocapsid (N), and analyzed by flow cytometry for cytokine production. **B**, Comparisons of effector T cells in each group - percentages of CD4+ or CD8+ cells, producing INFγ, TNFα or IL-17 in response to stimulation by each peptide pool. Each dot represents a patient, color coded: purple for children; green for adults with mild disease and red for adults with severe disease. All analyses are Kruskal-Wallis tests, and the p values are indicated over brackets. Significant differences are indicated by p values in a higher font.

**Figure 6.**
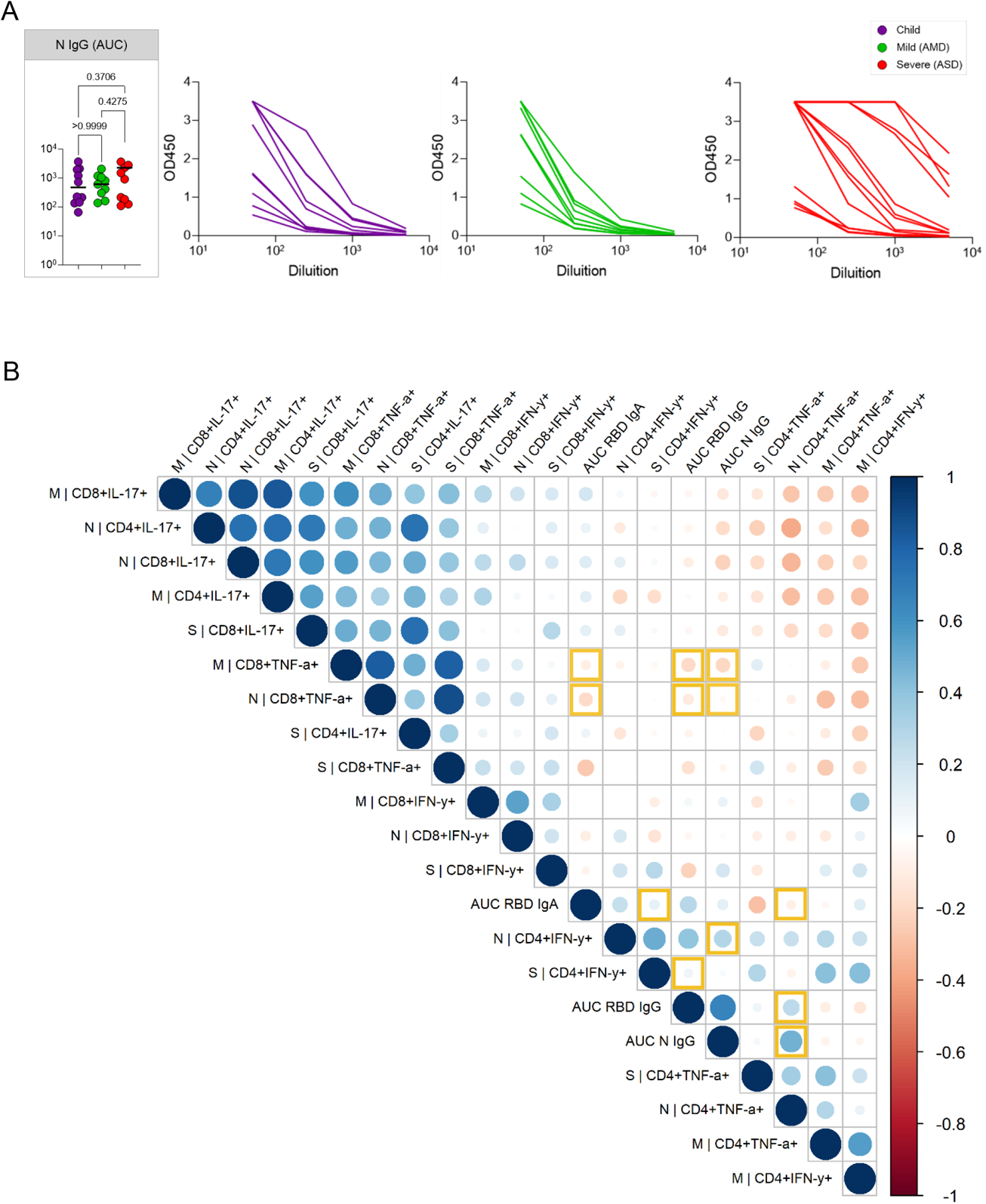
Anti-N IgG response and correlation of specific responses. **A**, SARS-CoV-2 anti-N IgG antibody titers determined by ELISA using serial dilutions of plasma. Individual titration curves for each individual (represented by a line, color coded) and analysis of variance (Kruskal-Wallis) of the values calculated as the area under the curve (AUC) for IgG (**A**) are displayed. P values are displayed over brackets. **B**, Matrix representing a Spearman correlation analysis of specific effector T cells responses (in percentages of positive CD4+ and CD8+ positive cytokine expressing cells in response to peptide pools) and the antibody response to the RBD of the spike protein and to the N protein (represented as values for the AUC). Correlations between the specific effector CD8+ T and CD4+ T cells frequencies, and the antibody (AUC – area under the curve) values for the RBD and N protein are highlighted.

## Discussion

It is clear from our study as well as from others (23) children do get infected by SARS-CoV-2, and thus possibly contribute to the community-based spread of the virus, contrary to what has been suggested by studies on the low nasal ACE2 expression in children (24). The lower rates of infection in children can be biased by lower testing, as suggested by (25), and should be more carefully studied, given its importance for planning school openings. Our findings on the more naïve, non-specific lymphocyte profile presented by in children, if taken isolated from the other results in this study, could indicate that a naive immune system does better than an old one, as has been suggested (9) – that children would be better equipped to mount fast and efficient immune responses to rapidly clear the virus. However, one prediction from this hypothesis is that children would be more likely to present mild forms of all viral diseases, and this is not the case. While milder manifestations are observed in MERS, SARS, and varicella, the opposite is observed for infection with poliovirus, and also respiratory viruses, especially influenza and respiratory syncytial virus (RSV) - reviewed in (6).

Our data indicate the possibility that not only the adaptive but also their innate immune system has relevant characteristics that enabled children to mount an efficient immune response and control the infection. The important differences observed for dendritic cells might offer an important clue. DCs play crucial roles in initiating and shaping the adaptive response, and subpopulations of DCs, especially pDC, are determinants for the generation of efficient antiviral responses, being one of the main sources of type I interferon (26). A previous study in COVID-19 adult patients indicated decreased activation and numbers of DCs (27). In our study, children consistently showed higher frequencies of DCs, including pDCs, compared to adults. High HLA-DR expression is characteristic of mature DCs; as they become activated by engagement of pattern recognizing receptors, HLA-DR first increases, and then decreases as DCs migrate to draining lymph nodes (26,28) Low HLA-DR expression in children is associated with immune suppression (4) and acute inflammatory conditions (29). The inverse correlation of HLA-DR with CX3CR1 in DCs is intriguing. CX3CR1, also known as the fractalkine receptor, is considered a homing marker for inflamed tissue and plays a role in pathology in Japanese virus-induced encephalitis ref (30) and peritoneal vasculitis in a sepsis model (31). The high levels of HLA-DR in children’s DCs indicate that their cells are not poorly activated, but mature and able to generate efficient immune responses. The low expression of CX3CR1 suggests that children’s DCs are not targeted to inflamed sites, as they seem to be in ASD. Our results indicate that expression of CX3CR1 in circulating DCs could associate, or serve as a marker for, pathological mechanisms in severe COVID-19, and suggest that inflammation to specific sites such as the lung may be affected by age.

Pediatric patients in our sample presented SARS-CoV-2-specific antibodies and T cell responses in levels comparable to adult patients. Based on our data, we propose that the higher CD8+TNFα+ responses for the M and N proteins could be associated with a protective response in children. The M protein is the most abundant structural protein on the surface of the virus (32), thus it could potentially constitute an important target of immune responses. A study by Thieme et al. (33) found anti-M CD4+ T cells as the highest T cell response in critical COVID-19 patients. In that study, CD4+ T, rather than CD8+ T cell immunity to SARS-CoV-2 proteins dominated the response in severe and critical patients, indicating that a robust CD4+ T cell response to these antigens was not determinant for protection. The nucleocapsid (N) protein is structural, and though not expressed on the surface, abundantly produced upon infection, and highly conserved among beta coronaviruses (34). This protein was a major target for early B cell responses in the SARS epidemic of 2003 (35). More recent studies (17,36) found strong T cell responses to the N protein in SARS-CoV-2 patients, and also in individuals who recovered from SARS. These studies support a relevant role for structural N protein as an immune target in SARS-CoV-2 infection. We found robust antibody responses in children both to the S RBD as well as to the N protein, while Weisberg et al. (9) found antibodies to the S, but not the N protein; and Cohen et al. (37) found lower responses in children in general. These differences, as well as a lower response to the S protein in general in our sample, may reflect differences in HLA between American and Brazilian populations, or even a difference in immunization history, given a tradition in vaccination program for children in Brazil. The correlations of antibody responses with CD4+ T cells are somewhat expected, given the help needed for antibody production, and indicate that these responses are somewhat coordinated, but also that not all antibodies produced are linked to TNFα or INFγ help. Our next studies will focus on further characterizing this antibody response in detail. The inverse correlation between specific CD8+ T cell responses and antibodies may indicate a relevant role for cytotoxic immunity against SARS-CoV-2, beyond antibody production.

Children are considered the natural reservoir of seasonal coronaviruses that cause the common cold (4). A hypothesis frequently raised to explain milder disease in children with COVID-19 is that the presence of neutralizing antibodies to such viruses could cross-protect them upon infection with SARS-CoV-2. However, a recent study in adults found no evidence of cross-protection associated with levels of these antibodies (38). Alternatively, protection could be conferred not by cross-reactive antibodies, but rather by pre-existing N-protein-specific T cells. Most studies - and most vaccines - have so far focused on protection against SARS-CoV-2 infection by antibodies to the spike protein. Both screen studies by Ferreti et al. and Ng et al. indicated that T cell immunity to SARS-CoV-2 infected individuals includes many targets outside the spike protein and that they are not conserved among coronaviruses that cause the common cold. These findings are in agreement with the ones of the Le Bert study. Our results support that the role of T cell responses to the N protein must be further investigated, with a more detailed T cell epitope mapping. Such work might reveal additional correlates of protection, and/or epitopes to add in the next generation of COVID-19 vaccines. A recent report (39) indicated that T cell immunity was not markedly affected, so far, by the emergence of new variants, supporting the study of T cell epitopes to be added to the next vaccines.

The main limitation of this study is that it is mostly an exploratory, descriptive one, and compares individuals in different age groups. We believe this was a valid approach given the magnitude of what is unknown at the moment. Usually, biomarkers in peripheral blood are only useful when highly correlated with outcomes. Yet, most studies that seek to understand how immune responses can correlate with protection compare adult s with mild and severe disease, and it is known that these are in different age groups. At present, the best age-matched controls for SARS-CoV-2 infected patients are still unknown. Certainly, the absence of a pre-pandemic healthy children control group is a limitation of this and all the other studies that focused on the general, non-specific immune profile of children with COVID-19. There is still much to be understood about immunological differences not only between pediatric and adult COVID-19 patients but also in other diseases. In this sense, we believe our work contributes significant information which was collected in a completely unbiased investigation -we did not know what differences or similarities to expect. At the time the project started, COVID-19 numbers in Brazil were still not high and the frequency of MISC patients or children with severe manifestations of the disease was still too low to include. Inclusion of patients was easiest in ASD because they were admitted to the hospital, thus their time of symptoms to sample collection is shorter compared to AMD and children. However, we do not think this was a major influence in the results - if so, differences would have been highest between ASD and AMD, and that was not the case. Finally, these results are based on a single point of collection and we still do not know how they will evolve into the memory responses that will be ultimately generated. The study on this cohort is still ongoing, with two more points of sample collection. We expect that further analysis of our data, as well as other studies’, on immune profile data and specific responses, will bring relevant information on the generation of immune memory in pediatric COVID-19.

## Data Availability

Raw data may be available upon request.

## Acknowledgments

Funding for this study was provided by PROADI - HMV, and the Ministry of Health; fellowships for Karina Lima, Julia Fontoura, Renato Stein, and Cristina Bonorino are from CNPq; fellowships for Gabriel Hilario, Priscila Oliveira, and Tiago Fazolo are from CAPES. TJB is a recipient of an American Heart Association fellowship grant. We wish to thank Drs. André Báfica, Daniel Mansur, Helder Nakaya, Leo Riella, Graham Pawelec and Steve Hedrick for critical readings of this manuscript. Finally, we wish to thank all the patients who accepted to enroll in the study and donate blood.

## Tables

Supplementary Table 1. Principal Component Analysis (PCA) variances and loading.

## Supplementary figures

1. Gating strategies for the key cell populations.
2. Spearman correlation of all variables composing the immune profile.
3. Principal Component Analysis of adaptive cells immune signatures.
4. Analysis of variance (Kruskall-Wallis) of remaining immune variables.
5. Control gate strategies for flow cytometry analysis of specific T cell responses.
6. Comparison of specific T cell responses by effector T cell type among the groups.

