## Supplemental Data for "Strong anti-viral responses in pediatric COVID-19 patients in South Brazil"

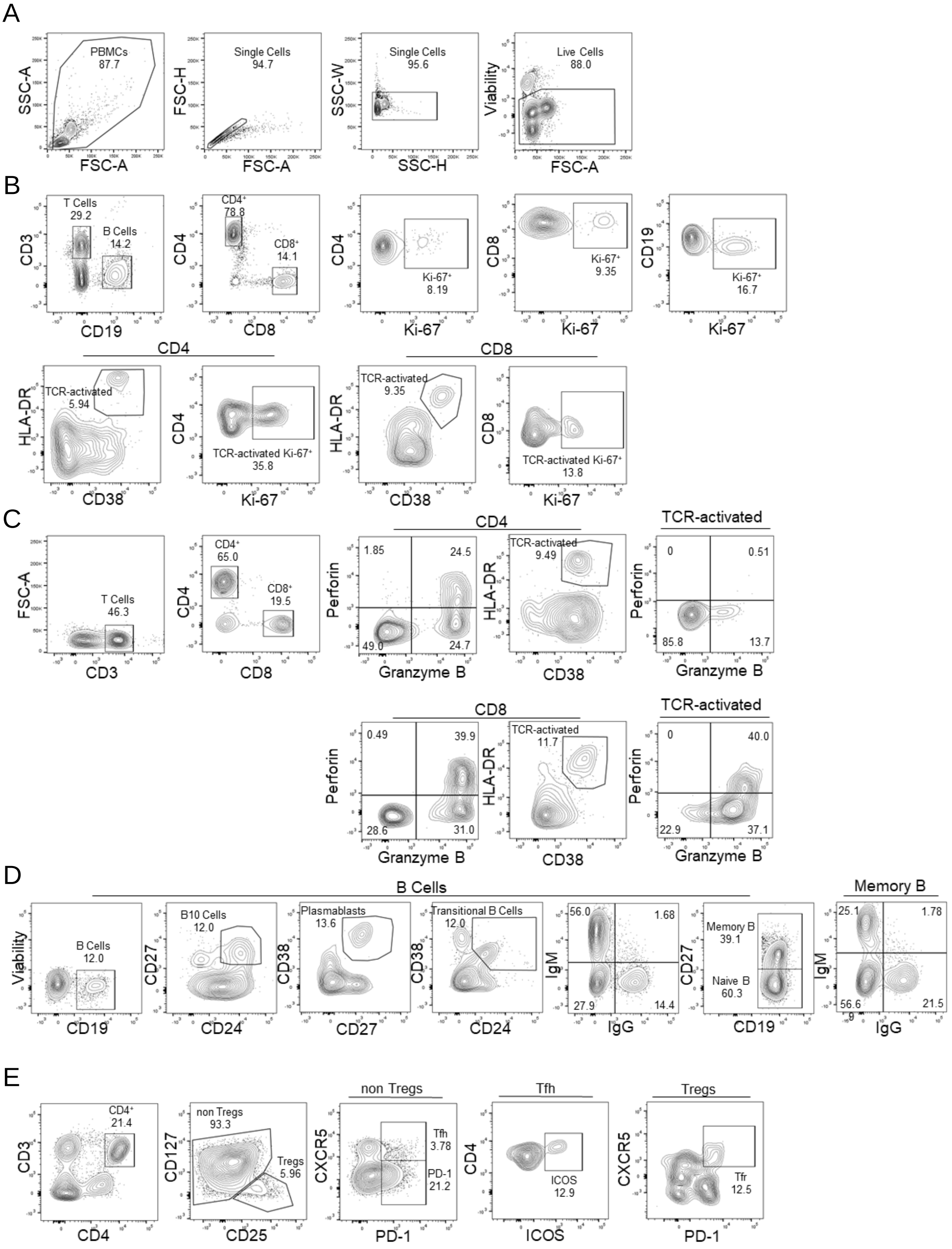
A
PBMCs 87.7
SSC-A
FSC-A
Single Cells 94.7
FSC-H
Single Cells 95.6
SSC-W
SSC-H
Live Cells 88.0
Viability
B
T Cells 29.2
B Cells 14.2
CD3
CD19
CD4+ 78.8
CD8+ 14.1
CD4
CD8
Ki-67+ 8.19
Ki-67
Ki-67+ 9.35
Ki-67+ 16.7
CD4
TCR-activated 5.94
HLA-DR
CD38
TCR-activated Ki-67+ 35.8
CD8
TCR-activated 9.35
TCR-activated Ki-67+ 13.8
C
T Cells 46.3
FSC-A
CD4+ 65.0
CD8+ 19.5
1.85
24.5
49.0
24.7
Perforin
Granzyme B
TCR-activated 9.49
TCR-activated
0
0.51
85.8
13.7
0.49
39.9
28.6
31.0
TCR-activated 11.7
0
40.0
22.9
37.1
D
B Cells
B Cells 12.0
Viability
B10 Cells 12.0
CD27
CD24
Plasmablasts 13.6
CD38
CD27
Transitional B Cells 12.0
56.0
1.68
27.9
14.4
IgM
IgG
Memory B 39.1
Naive B 60.3
Memory B
25.1
1.78
56.6
21.5
E
CD4+ 21.4
non Tregs 93.3
Tregs 5.96
CD127
CD25
non Tregs
Tfh 3.78
PD-1 21.2
CXCR5
PD-1
Tfh
ICOS 12.9
ICOS
Tregs
Tfr 12.5

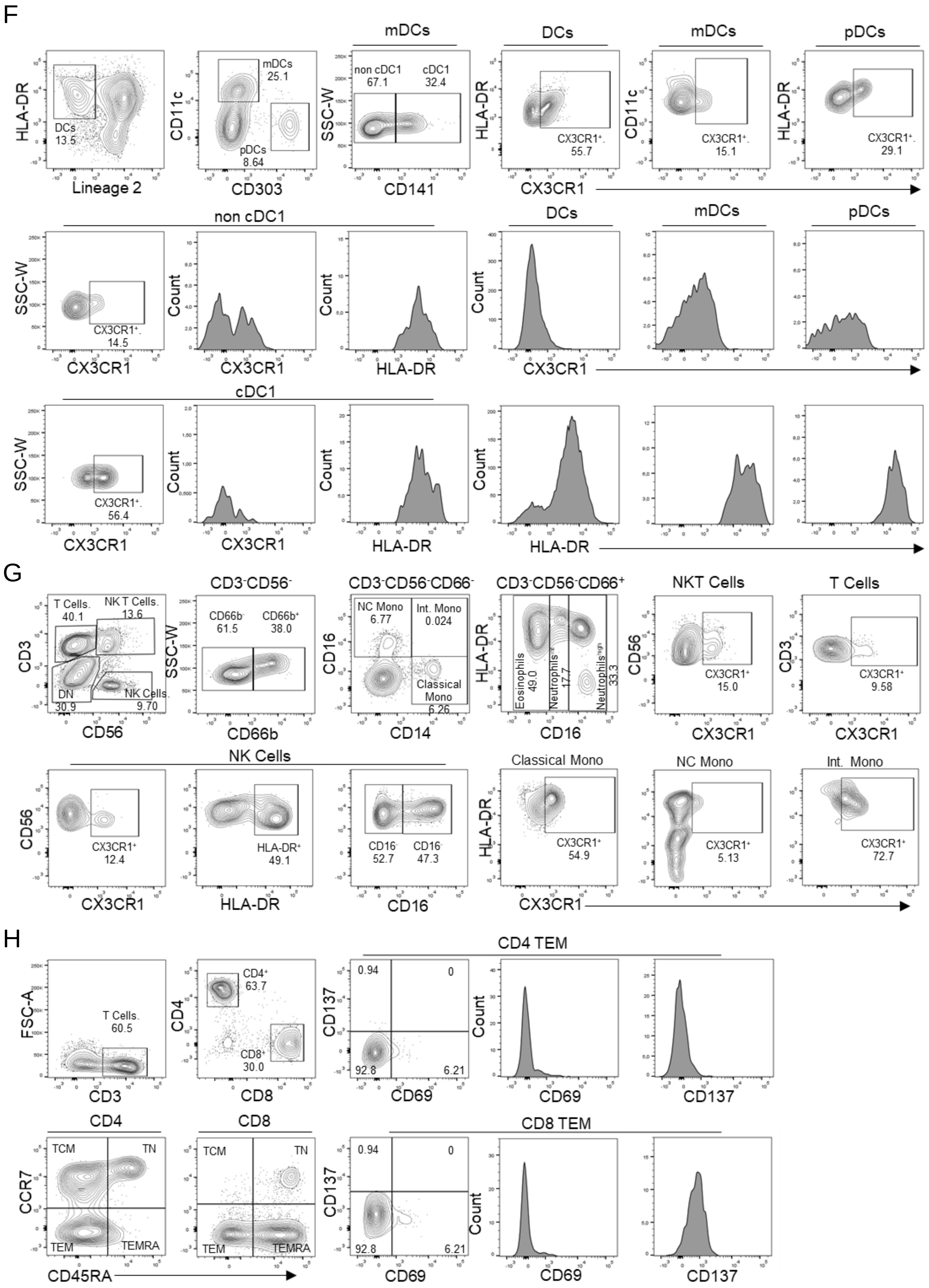

F
HLA-DR
DCs
13.5
Lineage 2
CD11c
mDCs
25.1
pDCs
8.64
CD303
mDCs
SSC-W
non cDC1
67.1
cDC1
32.4
CD141
DCs
HLA-DR
CX3CR1⁺
55.7
mDCs
CD11c
CX3CR1⁺
15.1
pDCs
HLA-DR
CX3CR1⁺
29.1
CX3CR1
non cDC1
SSC-W
CX3CR1⁺
14.5
CX3CR1
Count
CX3CR1
Count
HLA-DR
DCs
mDCs
pDCs
Count
CX3CR1
cDC1
SSC-W
CX3CR1⁺
56.4
CX3CR1
Count
CX3CR1
Count
HLA-DR
Count
HLA-DR
G
T Cells.
40.1
NK T Cells.
13.6
CD3
DN
30.9
NK Cells.
9.70
CD56
CD3⁻CD56⁻
CD66b⁻
61.5
CD66b⁺
38.0
SSC-W
CD66b
CD3⁻CD56⁻CD66⁻
NC Mono
6.77
Int. Mono
0.024
CD16
Classical Mono
6.26
CD14
CD3⁻CD56⁻CD66⁺
HLA-DR
Eosinophils
49.0
Neutrophils
17.7
Neutrophils
33.3
CD16
NKT Cells
CD56
CX3CR1⁺
15.0
CX3CR1
T Cells
CD3
CX3CR1⁺
9.58
CX3CR1
NK Cells
CD56
CX3CR1⁺
12.4
CX3CR1
HLA-DR⁺
49.1
HLA-DR
CD16⁻
52.7
CD16⁻
47.3
CD16
Classical Mono
HLA-DR
CX3CR1⁺
54.9
NC Mono
CX3CR1⁺
5.13
Int. Mono
CX3CR1⁺
72.7
CX3CR1
H
FSC-A
T Cells.
60.5
CD3
CD4
CD4⁺
63.7
CD8⁺
30.0
CD8
CD4 TEM
CD137
0.94
0
92.8
6.21
CD69
Count
CD69
CD137
CD4
CCR7
TCM
TN
TEM
TEMRA
CD45RA
CD8
TCM
TN
TEM
TEMRA
CD8 TEM
CD137
0.94
0
92.8
6.21
CD69
Count
CD69
CD137

**Supplementary Figure 1. Gating strategies for the key cell populations described in Fig. 1-4, Fig. 6A-B, Fig. 7, and other supplementary figures.** **A**, Gating strategy to identify all populations described in B-H. **B**, B and T cell surface staining gating strategy to identify B cells proliferation, CD4 and CD8 T cells, proliferation and TCR-activated T cells. **C**, T cell surface staining gating strategy to identify CD4 and CD8 T cells producing granzyme B and perforin in the population with or without TCR-activated T cells. **D**, B cell surface staining gating strategy to identify B10 cells, plasmablasts, transitional B cells, IgM cells, IgG cells, memory and naive cells populations. **E**, T cell surface staining gating strategy to identify Treg and TFH CD4 cells population. **F**, DCs surface staining gating strategy to identify differences in HLA-DR and CX3CR1 expression in DCs, mDC and pDCs cell populations. **G**, Innate cells and T surface staining gating strategy to identify NK cells, NK T cells, Eosinophils, Neutrophils and Monocytes populations. **H**, T cell surface staining gating strategy to identify differences in CD137 and CD69 expression in CD4 and CD8 T cells effector memory, central memory, terminally differentiated and naive population.

A

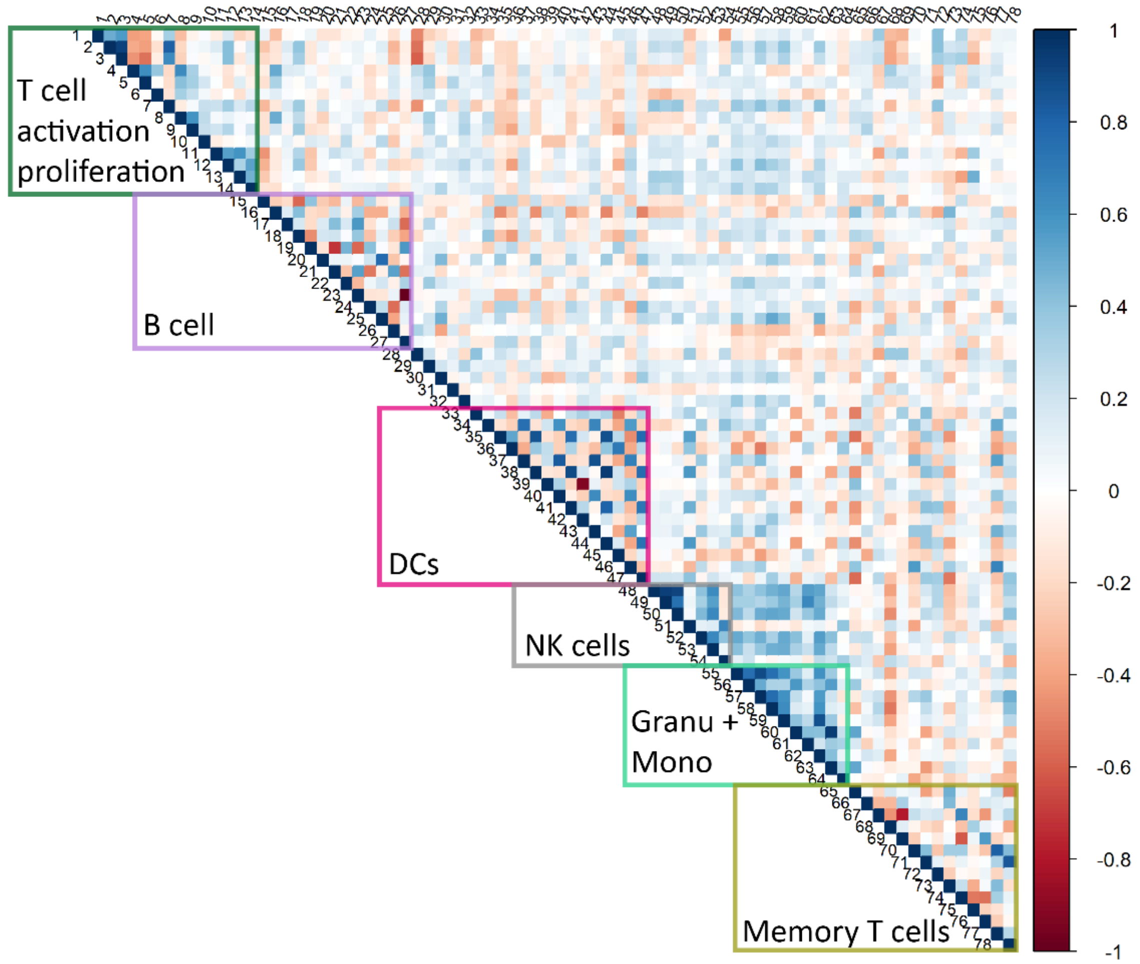


B

| | | | | | |
|---|---|---|---|---|---|
| 1 | PBMCs | 27 | Naive B Cells | 53 | HLA-DR$^+$ NK Cells |
| 2 | T cells | 28 | PD-1$^+$ CD4$^+$ T | 54 | CX3CR1$^+$ NK T cells |
| 3 | CD4$^+$ | 29 | Tfh cells | 55 | CD66$^+$ Granulocytes |
| 4 | Ki-67$^+$ CD4$^+$ | 30 | ICOS$^+$ Tfh cells | 56 | CD66b$^+$ CD16 hi Neutrophils |
| 5 | TCR-activated CD4$^+$ | 31 | Treg | 57 | CD66b$^+$ CD16 int Neutrophils |
| 6 | TCR-activated Ki-67$^+$ CD4$^+$ | 32 | CXCR5$^+$ Treg | 58 | CD66b$^+$ CD16 low Eosinophils |
| 7 | CD8$^+$ | 33 | DCs | 59 | Classical Monocytes |
| 8 | Ki-67$^+$ CD8$^+$ | 34 | CX3CR1 gMFI DCs | 60 | Intermediate Monocytes |
| 9 | TCR-activated CD8$^+$ | 35 | HLA-DR gMFI DCs | 61 | NC Monocytes |
| 10 | TCR-activated Ki-67$^+$ CD8$^+$ | 36 | mDCs | 62 | CX3CR1$^+$ Classical Monocytes |
| 11 | GranzB$^+$Perf$^+$ CD4$^+$ | 37 | CX3CR1 gMFI mDCs | 63 | CX3CR1$^+$ Intermediate Monocytes |
| 12 | TCR-activated GranzB$^+$Perf$^+$ CD4$^+$ | 38 | HLA-DR gMFI mDCs | 64 | CX3CR1$^+$ NC Monocytes |
| 13 | GranzB$^+$Perf$^+$ CD8$^+$ | 39 | cDC1 | 65 | CX3CR1$^+$ T cells |
| 14 | TCR-activated GranzB$^+$Perf$^+$ CD8$^+$ | 40 | CX3CR1 gMFI cDC1 | 66 | CD4$^+$ TCM |
| 15 | B cells | 41 | HLA-DR gMFI cDC1 | 67 | CD4$^+$ T Naive |
| 16 | Ki-67$^+$ B cells | 42 | Non cDC1 | 68 | CD4$^+$ TEMRA |
| 17 | B10 cells | 43 | CX3CR1 gMFI Non cDC1 | 69 | CD4$^+$ TEM |
| 18 | Plasmoblasts | 44 | HLA-DR gMFI Non cDC1 | 70 | CD69$^+$ gMFI CD4$^+$ TEM |
| 19 | IgG$^-$IgM$^+$ B cells | 45 | pDCs | 71 | CD137$^+$ gMFI CD4$^+$ TEM |
| 20 | IgG$^+$IgM$^+$ B cells | 46 | CX3CR1 gMFI pDCs | 72 | CD8$^+$ |
| 21 | IgG$^+$IgM- B cells | 47 | HLA-DR gMFI pDCs | 73 | CD8$^+$ TCM |
| 22 | Transitional B cells | 48 | NK cells | 74 | CD8$^+$ T Naive |
| 23 | Memory B cells | 49 | CD16$^+$NK cells | 75 | CD8$^+$ TEMRA |
| 24 | IgG$^-$IgM$^+$ Memory B cells | 50 | CD16$^-$ NK cells | 76 | CD8$^+$ TEM |
| 25 | IgG$^+$IgM$^+$ Memory B cells | 51 | NK T cells | 77 | CD69$^+$ gMFI CD8$^+$ TEM |
| 26 | IgG$^+$IgM$^-$ Memory B cells | 52 | CX3CR1$^+$ NK Cells | 78 | CD137$^+$ gMFI CD8$^+$ TEM |

**Supplementary Figure 2**. **Spearman correlation of all variables composing the immune profile**. **A**, Clusters of more correlated variables are outlined and identified in a Spearman correlation matrix of all variables. **B**, List of immune variables indicated by numbers by which they are plotted on the matrix.

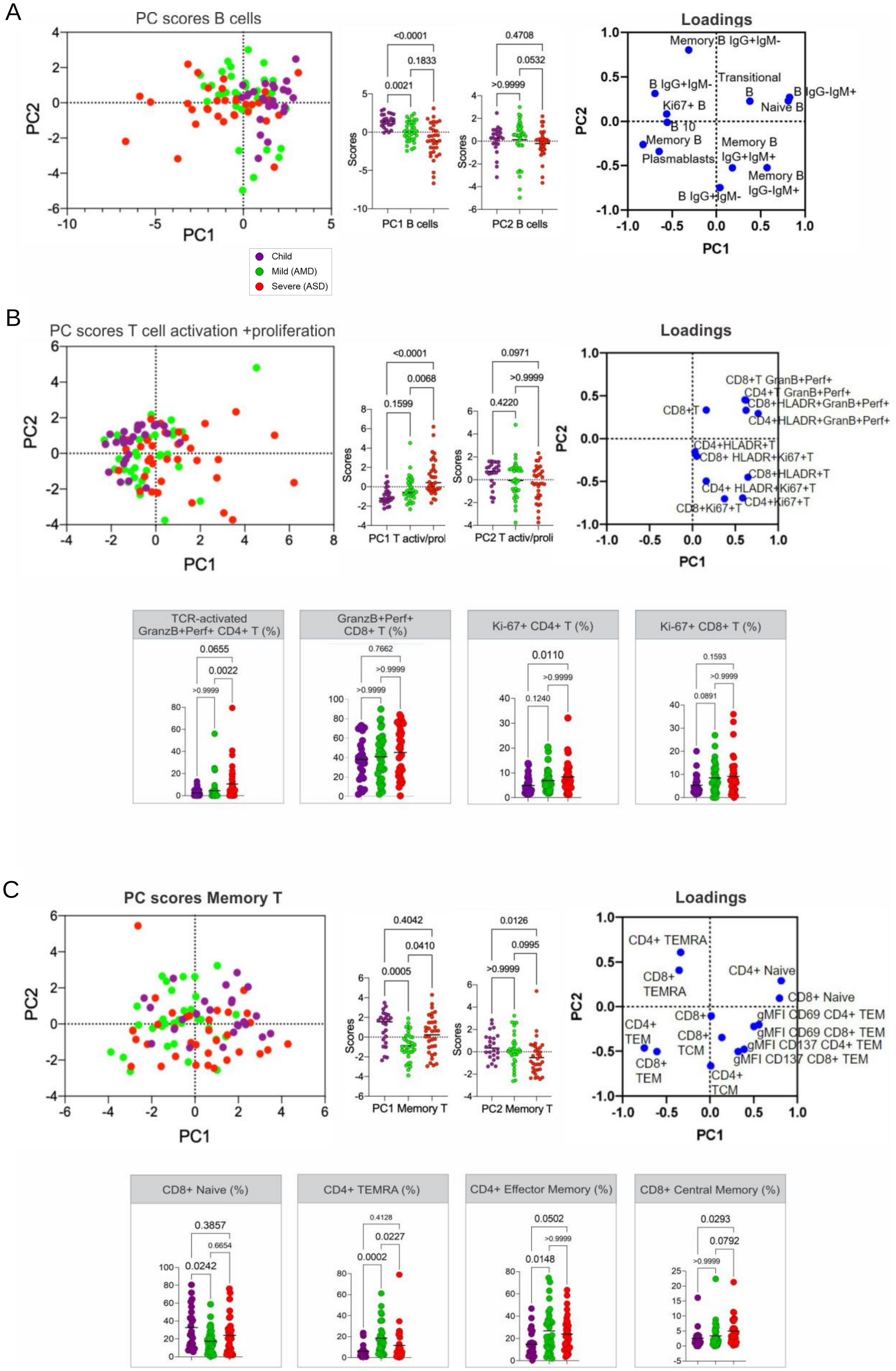
A
PC scores B cells
PC1
PC2
Child
Mild (AMD)
Severe (ASD)
<0.0001
0.1833
0.0021
Scores
PC1 B cells
0.4708
0.0532
>0.9999
PC2 B cells
Loadings
Memory B IgG+IgM-
Transitional B
B IgG+IgM-
B IgG-IgM+
Naive B
Ki67+ B
B 10
Memory B
Plasmablasts
Memory B IgG+IgM+
Memory B IgG-IgM+
B IgG+IgM-
B
PC scores T cell activation +proliferation
<0.0001
0.0068
0.1599
PC1 T activ/prol
0.0971
>0.9999
0.4220
PC2 T activ/proli
Loadings
CD8+T GranB+Perf+
CD4+T GranB+Perf+
CD8+HLADR+GranB+Perf+
CD4+HLADR+GranB+Perf+
CD8+T
CD4+HLADR+T
CD8+ HLADR+Ki67+T
CD8+HLADR+T
CD4+ HLADR+Ki67+T
CD8+Ki67+T
CD4+Ki67+T
TCR-activated GranzB+Perf+ CD4+ T (%)
0.0655
0.0022
>0.9999
GranzB+Perf+ CD8+ T (%)
0.7662
>0.9999
>0.9999
Ki-67+ CD4+ T (%)
0.0110
>0.9999
0.1240
Ki-67+ CD8+ T (%)
0.1593
>0.9999
0.0891
C
PC scores Memory T
0.4042
0.0410
0.0005
PC1 Memory T
0.0126
0.0995
>0.9999
PC2 Memory T
Loadings
CD4+ TEMRA
CD8+ TEMRA
CD4+ Naive
CD8+ Naive
gMFI CD69 CD4+ TEM
gMFI CD69 CD8+ TEM
gMFI CD137 CD4+ TEM
gMFI CD137 CD8+ TEM
CD4+ TEM
CD8+ TEM
CD8+
CD8+ TCM
CD4+ TCM
CD8+ Naive (%)
0.3857
0.6654
0.0242
CD4+ TEMRA (%)
0.4128
0.0227
0.0002
CD4+ Effector Memory (%)
0.0502
>0.9999
0.0148
CD8+ Central Memory (%)
0.0293
0.0792
>0.9999

**Supplementary Figure 3. Principal Component Analysis of adaptive cells immune signatures. A**, B cells; **B**, Proliferating/activated T cells; and **C**, Memory T cells). For each signature, are displayed the PCA plot of PC1xPC2, the differences in scores of individuals for each PC; the loadings of the main variables contributing to each PC; and Kruskal-Wallis tests comparisons of the major contributing variables values for each group of patients.

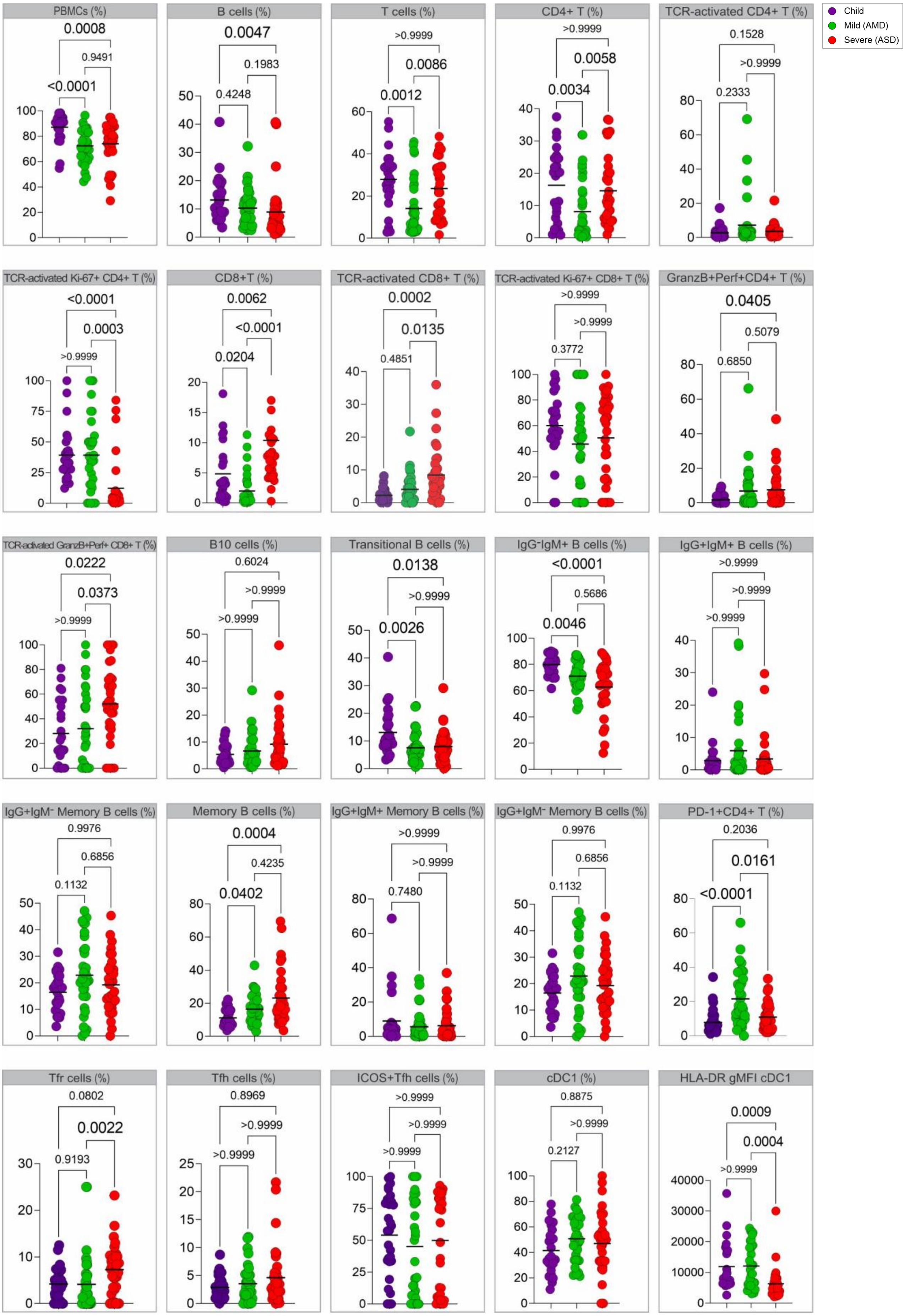

Child
Mild (AMD)
Severe (ASD)
PBMCs (%)
0.0008
0.9491
<0.0001
B cells (%)
0.0047
0.1983
0.4248
T cells (%)
>0.9999
0.0086
0.0012
CD4+ T (%)
>0.9999
0.0058
0.0034
TCR-activated CD4+ T (%)
0.1528
>0.9999
0.2333
TCR-activated Ki-67+ CD4+ T (%)
<0.0001
0.0003
>0.9999
CD8+T (%)
0.0062
<0.0001
0.0204
TCR-activated CD8+ T (%)
0.0002
0.0135
0.4851
TCR-activated Ki-67+ CD8+ T (%)
>0.9999
>0.9999
0.3772
GranzB+Perf+CD4+ T (%)
0.0405
0.5079
0.6850
TCR-activated GranzB+Perf+ CD8+ T (%)
0.0222
0.0373
>0.9999
B10 cells (%)
0.6024
>0.9999
>0.9999
Transitional B cells (%)
0.0138
>0.9999
0.0026
IgG⁻IgM+ B cells (%)
<0.0001
0.5686
0.0046
IgG+IgM+ B cells (%)
>0.9999
>0.9999
>0.9999
IgG+IgM⁻ Memory B cells (%)
0.9976
0.6856
0.1132
Memory B cells (%)
0.0004
0.4235
0.0402
IgG+IgM+ Memory B cells (%)
>0.9999
>0.9999
0.7480
IgG+IgM⁻ Memory B cells (%)
0.9976
0.6856
0.1132
PD-1+CD4+ T (%)
0.2036
0.0161
<0.0001
Tfr cells (%)
0.0802
0.0022
0.9193
Tfh cells (%)
0.8969
>0.9999
>0.9999
ICOS+Tfh cells (%)
>0.9999
>0.9999
>0.9999
cDC1 (%)
0.8875
>0.9999
0.2127
HLA-DR gMFI cDC1
0.0009
0.0004
>0.9999

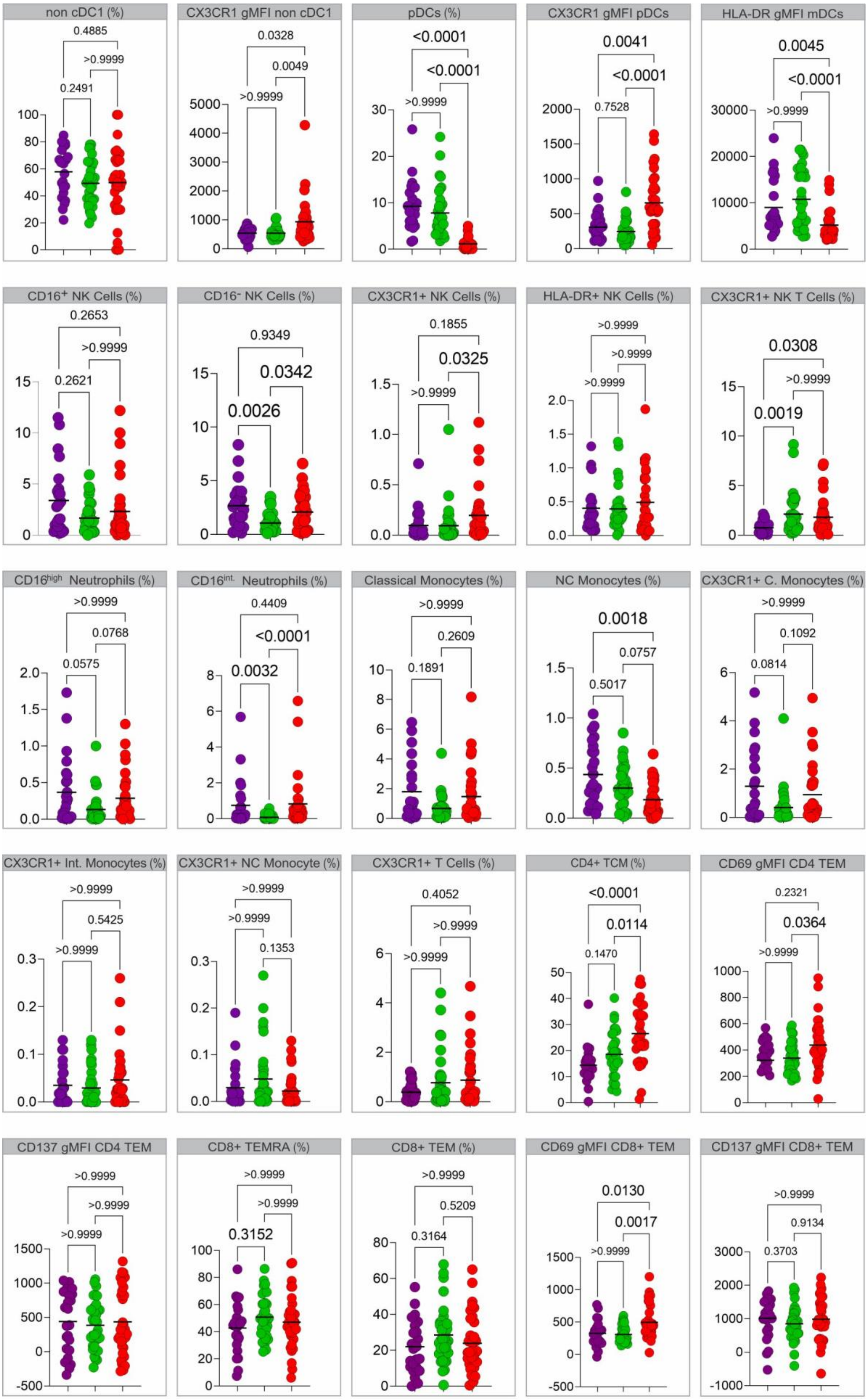
non cDC1 (%)
0.4885
>0.9999
0.2491
CX3CR1 gMFI non cDC1
0.0328
0.0049
>0.9999
pDCs (%)
<0.0001
<0.0001
>0.9999
CX3CR1 gMFI pDCs
0.0041
<0.0001
0.7528
HLA-DR gMFI mDCs
0.0045
<0.0001
>0.9999
CD16+ NK Cells (%)
0.2653
>0.9999
0.2621
CD16- NK Cells (%)
0.9349
0.0342
0.0026
CX3CR1+ NK Cells (%)
0.1855
0.0325
>0.9999
HLA-DR+ NK Cells (%)
>0.9999
>0.9999
>0.9999
CX3CR1+ NK T Cells (%)
0.0308
>0.9999
0.0019
CD16high Neutrophils (%)
>0.9999
0.0768
0.0575
CD16int. Neutrophils (%)
0.4409
<0.0001
0.0032
Classical Monocytes (%)
>0.9999
0.2609
0.1891
NC Monocytes (%)
0.0018
0.0757
0.5017
CX3CR1+ C. Monocytes (%)
>0.9999
0.1092
0.0814
CX3CR1+ Int. Monocytes (%)
>0.9999
0.5425
>0.9999
CX3CR1+ NC Monocyte (%)
>0.9999
>0.9999
0.1353
CX3CR1+ T Cells (%)
0.4052
>0.9999
>0.9999
CD4+ TCM (%)
<0.0001
0.0114
0.1470
CD69 gMFI CD4 TEM
0.2321
0.0364
>0.9999
CD137 gMFI CD4 TEM
>0.9999
>0.9999
>0.9999
CD8+ TEMRA (%)
>0.9999
>0.9999
0.3152
CD8+ TEM (%)
>0.9999
0.5209
0.3164
CD69 gMFI CD8+ TEM
0.0130
0.0017
>0.9999
CD137 gMFI CD8+ TEM
>0.9999
0.9134
0.3703

**Supplementary Figure 4**. **Analysis of variance (Kruskall-Wallis) of remaining immune variables.** Analysis of variance of the values for immune variables (in percentages or gMFI) that were lesser influencers of the three first principal components and thus not included in the main figures.

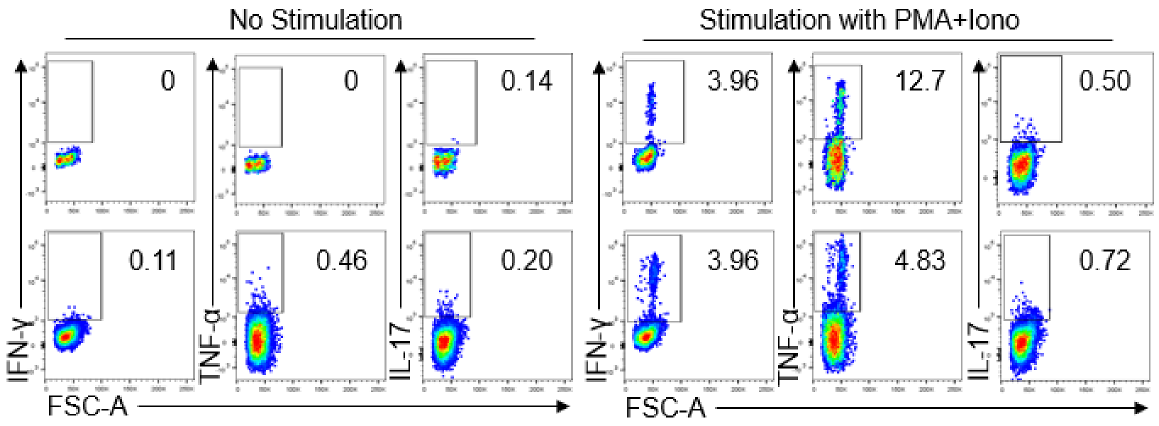


**Supplementary Figure 5**. **Control gate strategies for flow cytometry analysis of specific T cell responses.** Negative (DMSO) and positive (PMA+Ionomycin) controls gate strategies and representative plots of CD4+ or CD8+ T cell simulations.

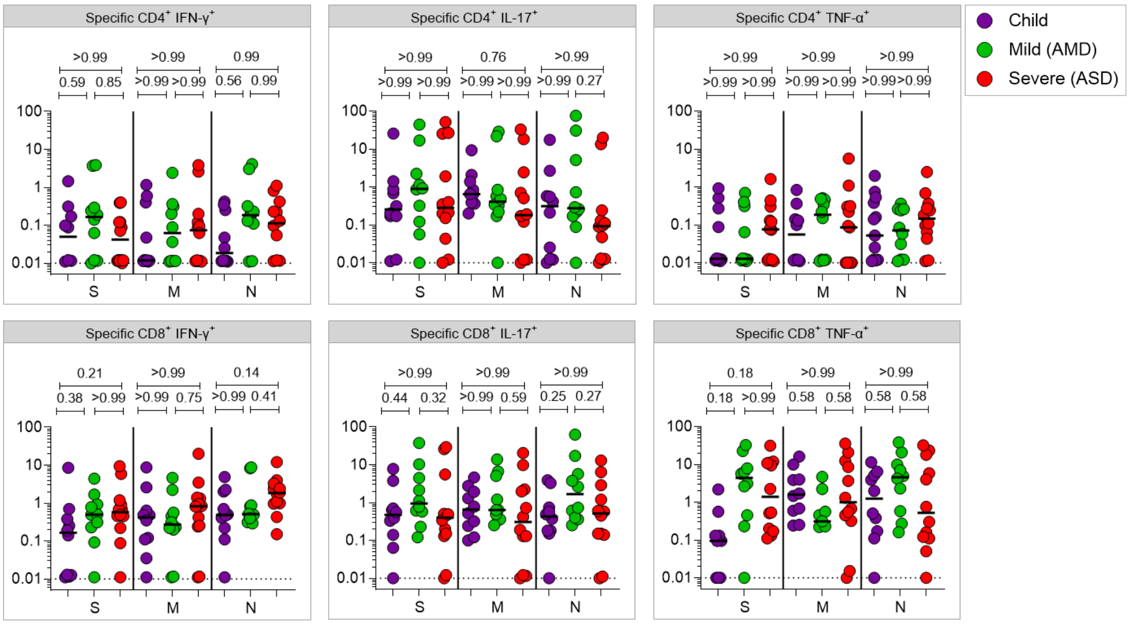


**Supplementary Figure 6**. **Comparison of specific T cell responses by effector T cell type among the groups.** Values of specific T cell responses (in percentages of positive CD4+ and CD8+ positive cytokine expressing cells in response to peptide pools) are plotted. Each dot represents a patient, color coded: children – purple, adult with mild disease – green, and adult with severe disease – red. All analyses are Kruskal-Wallis tests, and the p values are indicated in brackets.

**Supplementary Table 1.** Principal Component Analysis (PCA) variances and loading.

| **PCA all variables** | | | | | | | | | | | | | |
|---|---|---|---|---|---|---|---|---|---|---|---|---|---|
| Positive (loadings) | **PC1** | 1 | 2 | 3 | **PC2** | 1 | 2 | 3 | **PC3** | 1 | 2 | 3 | |
| | | Memory B IgG-IgM+ | Naïve B cells | cDC1 gMFI DR | | Treg | mDC | CD4+ TEMRA | | Naïve CD8+T | Naïve CD4+T | pDC | %cumul variance |
| | | 0.57 | 0.56 | 0.55 | | 0.53 | 0.45 | 0.40 | | 0.56 | 0.43 | 0.35 | |
| % variance explained | 12.16 | | | | 8.53 | | | | 8.12 | | | | 28.81 |
| Negative (loadings) | **PC1** | 1 | 2 | 3 | **PC2** | 1 | 2 | 3 | **PC3** | 1 | 2 | 3 | |
| | | B cells Ki67+ | Plasma blasts | DC gMFI CX3CR1 | | Eosino phils | NK cells | Granulo cytes | | DCs | mDC gMFI DR | cDC1 gMFI DR | |
| | | -0.71 | -0.58 | -0.60 | | -0.58 | -0.55 | -0.48 | | -0.64 | -0.60 | -0.60 | |

| **PCA Granu+Mono** | | | | | | | | |
|---|---|---|---|---|---|---|---|---|
| Positive (loadings) | **PC1** | 1 | 2 | 3 | **PC2** | 1 | 2 | |
| | | Classical Monocytes | Intermediate Monocytes | Intermediate Monocytes CX3CR1+ | | Classical Monocytes CX3CR1+ | CD16 Int Neutrophils | %cumul variance |
| | | 0.84 | 0.80 | 0.75 | | 0.43 | 0.37 | |
| % variance explained | 33.41 | | | | 16.96 | | | 50.37 |
| Negative (loadings) | **PC1** | 1 | 2 | 2 | **PC2** | 1 | 2 | |
| | | - | - | - | | NC Monocytes | NC Monocytes CX3CR1+ | |
| | | - | - | - | | -0.66 | -0.64 | |

| **PCA NK cells** | | | | | | | | |
|---|---|---|---|---|---|---|---|---|
| Positive (loadings) | **PC1** | 1 | 2 | 3 | **PC2** | 1 | 2 | |
| | | NK Cells HLA-DR+ | NK Cells HLA-DR+ | NK CX3CR1+ | | NK T CX3CR1+ | NK T | %cumul variance |
| | | 0.85 | 0.77 | 0.73 | | 0.85 | 1.87 | |
| % variance explained | 37.42 | | | | 29.94 | | | 67.35 |
| Negative (loadings) | **PC1** | 1 | 2 | 3 | **PC2** | 1 | 2 | |
| | | - | - | - | | NK Cytotoxic | NK Regulatory | |
| | | - | - | - | | -0.43 | -0.34 | |

| **PCA Dendritic Cells** | | | | | | | | | |
|---|---|---|---|---|---|---|---|---|---|
| Positive (loadings) | **PC1** | 1 | 2 | 3 | **PC2** | 1 | 2 | 3 | |
| | | mDCs gMFI CX3CR1 | cDC1 gMFI CX3CR1 | Non cDC1 gMFI CX3CR1 | | pDCs | mDCs | cDC1 | % cumul variance |
| | | 0.57 | 0.51 | 0.44 | | 0.59 | 0.57 | 0.23 | |

| | | | | | | | | | |
|---|---|---|---|---|---|---|---|---|---|
| % variance explained | 35.81 | | | | 27.36 | | | | 63.17 |
| Negative (loadings) | **PC1** | 1 | 2 | 3 | **PC2** | 1 | 2 | 3 | |
| | | mDCs gMFI DR | DCs gMFI DR | cDC1 gMFI DR | | cDC1 gMFI CX3CR1 | Non cDC1 gMFI CX3CR1 | cDC1 gMFI DR | |
| | | -0.85 | -0.85 | -0.83 | | -0.60 | -0.53 | -0.49 | |
| **PCA T cell activation + proliferation** | | | | | | | | | |
| Positive (loadings) | **PC1** | 1 | 2 | 3 | **PC2** | 1 | 2 | | % cumul variance |
| | | CD4+ T HLADR+ GranB+Perf+ | CD8+ T HLADR+ | - | | CD4 T Ki67+ | CD8 T Ki67+ | | |
| | | 0.76 | 0.64 | - | | 0.55 | 0.43 | | |
| % variance explained | 24.22 | | | | 19.97 | | | | 44.19 |
| Negative (loadings) | **PC1** | 1 | 2 | 3 | **PC2** | 1 | 2 | | |
| | | CD8+ T Ki-67+ | CD4+ T Ki67+ | CD4+ T HLADR+ GranB+ Perf+ | | CD4+ T GranB+ Perf+ | CD8+ T GranB+ Perf+ | | |
| | | -0.70 | -0.67 | -0.59 | | -0.64 | -0.54 | | |
| **PCA B cells** | | | | | | | | | |
| Positive (loadings) | **PC1** | 1 | 2 | 3 | **PC2** | 1 | 2 | | % cumul variance |
| | | B IgG- IgM+ | Naive B Cells | Memory B IgG- IgM+ | | Memory B IgG+ IgM- | - | | |
| | | 0.82 | 0.69 | 0.67 | | 0.75 | - | | |
| % variance explained | 31.2 | | | | 18.36 | | | | 49.56 |
| Negative (loadings) | **PC1** | 1 | 2 | 3 | **PC2** | 1 | 2 | | |
| | | B IgG+ IgM- | Plasmablasts | B cells Ki-67+ | | B IgG+ IgM+ | Memory IgG+ IgM+ | | |
| | | -0.75 | -0.67 | -0.59 | | -0.80 | -0.57 | | |
| **PCA T cell memory** | | | | | | | | | |
| Positive (loadings) | **PC1** | 1 | 2 | | **PC2** | 1 | 2 | | % cumul variance |
| | | CD4+ T Naive | CD8+ T Naive | | | CD4+ TEMRA | CD8+ TEMRA | | |
| | | 0.81 | 0.79 | | | 0.61 | 0.41 | | |
| % variance explained | 25.38 | | | | 17.1 | | | | 42.49 |
| Negative (loadings) | **PC1** | 1 | 2 | | **PC2** | 1 | 2 | | |
| | | CD4+ TEM | CD8+ TEM | | | CD4+ TCM | CD8+ TEM | | |
| | | -0.76 | -0.61 | | | -0.66 | -0.50 | | |
